# Whole Exome Sequencing of Nigerian Prostate Tumors from the Prostate Cancer Transatlantic Consortium (CaPTC) Reveals DNA Repair Genes Associated with African Ancestry

**DOI:** 10.1101/2022.02.17.22271157

**Authors:** Jason White, Ernest T Kaninjing, Kayode A Adeniji, Paul Jibrin, John O Obafunwa, Chidiebere N Ogo, Mohammed Faruk, Ademola Popoola, Omolara A Fatiregun, Olabode P Oluwole, Balasubramanyam Karanam, Isra Elhussin, Stefan Ambs, Wei Tang, Melissa Davis, Paz Polak, Moray J Campbell, Damian K Francis, Denise Y Gibbs, Kathryn R Brignole, Solomon Rotimi, Folake T Odedina, Damali N Martin, Clayton Yates

## Abstract

In this study, we used whole-exome sequencing of a cohort of 45 advanced-stage, treatment-naïve Nigerian (NG) primary prostate cancer (PCa) tumors and 11 unmatched non-tumor tissues to compare genomic alterations with African American (AA) and European American (EA) TCGA PCa. NG samples were collected from 6 sites in central and southwest Nigeria. After whole-exome sequencing, samples were processed using GATK best practices. *BRCA1* (100%), *BARD1* (45%), *BRCA2* (27%), and *PMS2*(18%) had germline alterations in at least two NG non-tumor samples. Across 111 germline variants, the AA cohort reflected a pattern [*BRCA1* (68%), *BARD1* (34%), *BRCA2* (28%), and *PMS2* (16%)] similar to NG samples. Of the most frequently mutated genes, *BRCA1* showed a statistically (p ≤ 0.05) higher germline mutation frequency in men of African ancestry (MAA) and increasing variant frequency with increased African ancestry. Disaggregating gene level mutation frequencies by variants revealed both ancestry-linked and NG-specific germline variant patterns. Driven by rs799917 (T>C), *BRCA1* showed an increasing mutation frequency as African admixture increased. BRCA2_rs11571831 was present only in MAAs, and BRCA2_rs766173 was elevated in NG men. 133 somatic variants were present in 26 PCa-associated genes within the NG tumor cohort. *BRCA2* (27%), *APC* (20%), *ATM* (20%), *BRCA1* (13%), *DNAJC6* (13%), *EGFR* (13%), *MAD1L1* (13%), *MLH1* (11%), and *PMS2* (11%) showed mutation frequencies > 10%. Compared to TCGA cohorts, NG tumors showed statistically significant elevated frequencies of *BRCA2, APC*, and *BRCA1*. The NG cohort variant pattern shared similarities (cosign similarities ≥ 0.734) with COSMIC signatures 5 and 6, and mutated genes showed significant (q < 0.001) GO and functional enrichment in mismatch repair and non-homologous repair deficiency pathways. Here, we showed that alterations in DNA damage response (DDR) genes were higher in NG PCa samples and that a portion of those alterations correlate with African ancestry. Moreover, we identified variants of unknown significance that may contribute to population-specific routes of tumorigenesis and treatment. These results present the most comprehensive characterization of the NG PCa exome to date and highlight the need to increase diversity of study populations.

## Introduction

For men, adenocarcinoma of the prostate is the most frequently diagnosed cancer, accounting, globally, for an estimated 1,414,259 cases and 375,304 deaths in 2020. The preponderance of this mortality is for men of African ancestry (MAA), including African American (AA), Central American, Caribbean, and Sub-Saharan African men^1^. The Globocan 2020-derived mortality-to-incidence ratio (M/I), “an indirect description of the general survival experience” of prostate cancer (PCa) in Africa, is 0.55, relative to 0.32 and 0.17 for Asia and Europe, respectively^2,3^. Furthermore, studies of AA men showed higher incidence, worse prognoses, and higher mortality compared to European American (EA) men^4,5^. Although there is a substantial contribution of social and environmental influence on the disparity, emerging evidence from genomic profiles suggests that this disease is highly heterogenous^6^, and its etiology and phenotype are influenced by enrichment of African ancestral genetic markers, with West African ancestry linked with higher Gleason grade at diagnosis^7,8^.

The racial disparity in PCa biology is typically characterized by increased genomic alterations, resulting in a more aggressive phenotype. For instance, West African ancestry is associated with distinctive somatic genomic alterations^9,10^. Conversely, understanding these putatively targetable genomic alterations presents opportunities for effective population-relevant and genomics-guided interventions that can improve clinical outcomes. This relies on the availability of genomic data for tumors from MAA. However, despite an upsurge in genomic data for human cancers, the data on PCa from African sources are grossly underrepresented in the literature and genomics databases. For instance, AA samples account for only about 10% of The Cancer Genome Atlas (TCGA) PCa sample cohorts^9,11^. Consequently, this gross underrepresentation impedes deciphering of clinically actionable genomic alterations that could be used to develop precision interventions for MAA. Hence, it is imperative to increase the representation by sequencing the tumor genome of PCa in MAA.

Although Black men in the Americas generally have ancestral roots in the Atlantic coasts of Africa^12-14^, the translational impact of genomics data of AA men to indigenous Africans is limited due to the varying degree of Afro-European and intra-African admixtures of American Blacks^15,16^. Such admixture and variation in germline alterations influence gene expression and phenotype^17^; and are limiting factors in the understanding of the contribution of genetics to health disparities^18^. Hence, studying the genomic architecture of PCa in the indigenous African population is essential for advancing understanding of the contribution of African genetics to PCa biology and the phenotype of this disease in the African Diaspora. To date, inadequate attention has been given to generation of genomics studies of PCa among indigenous Africans. Aside from three genome-wide association studies of PCa in Ghanaian^19^, Ugandan^20^, and South African^21^ men, only Jaratlerdsiri et al.^22^ have reported whole-genome sequence data on tumors from indigenous sub-Saharan Africans. Their analysis of PCa of six South African Black men identified distinctive and elevated oncogenic driver mutations, with a high proportion of these recurrent mutations appearing early in tumorigenesis. They also showed that tumors of the African men they studied had fewer complex genomic rearrangements, loss of PTEN, and absent ERG fusions and PIK3CA mutations relative to AAs. Furthermore, apart from large deletions within the *BRCA2, DEFA1B*, and *MFF* genes, they did not report any pathogenic mutations in high-penetrance genes, such as *BRCA1, BRCA2, ATM*, and *CHK2* among the South African cohort ^22,23^. Previous studies have identified, for PCa of AAs, a high burden of mutations in these DNA repair genes^24,25^; suggesting that poly(adenosine diphosphate-ribose) polymerase (PARP) inhibitors could improve clinical outcomes for men of African ancestry with PCa^26^. The South African study is limited by the small sample size. Furthermore, the differences observed could be due to the low contribution of South-African Khoe-San ancient ancestry genes to the AA genetic pool, which is a source of bias^15,16^. It is therefore our hypothesis that genomic profiling of PCa in indigenous West African men will identify clinically actionable targets for precision intervention for MAA. The utility of clinical mutational profiling necessitates greater emphasis on identifying variants that can be used to help these underrepresented patients.

The primary aim of the present study was to analyze whole-exome sequencing (WES) of 45 Nigerian (NG) primary treatment-naive formalin-fixed, paraffin-embedded (FFPE) samples of PCa collected within the Prostate Cancer Transatlantic Consortium (CaPTC). Study of PCa of NG men allowed us to provide genetic information of the indigenous West Africa population with the highest genetic contribution to AA men^15^ and provide opportunities to investigate the shared genetic background of both groups for causal disease variants. As such, our data will be relevant for deriving actionable clinical information for PCa intervention for MAA.

## Methods

### Sample Collection and Genomic DNA Extraction

This study utilized 45 FFPE advanced-stage, treatment-naïve primary PCa samples collected from four participating clinical sites within the CaPTC network (Figure 1A). Before use, all samples were reviewed and approved by Institutional Review Boards of their respective clinical institutions in Nigeria (NG) and by the Institutional Review Board at Tuskegee University. Five 10 µm-thick curls were obtained from each block with >50% tumor and ≤50% necrosis and shipped to Q2 Solutions (Morrisville, NC) for DNA extraction and quality analysis. Following the manufacturer’s protocol, genomic DNA and total RNA were purified using Allprep DNA/RNA FFPE kits (Qiagen, Hilden, Germany). DNA quality and quantity were checked with Qubit 2.0 fluorometry (Life Technologies, Carlsbad, CA) and with KAPA hgDNA quantification and QC kits (Kappa Biosystems [Roche], Basel, Switzerland). DNA quality and quantity thresholds were >0.2 µg and a Q129/Q41 ratio >0.00225, respectively.

**Figure 1.**
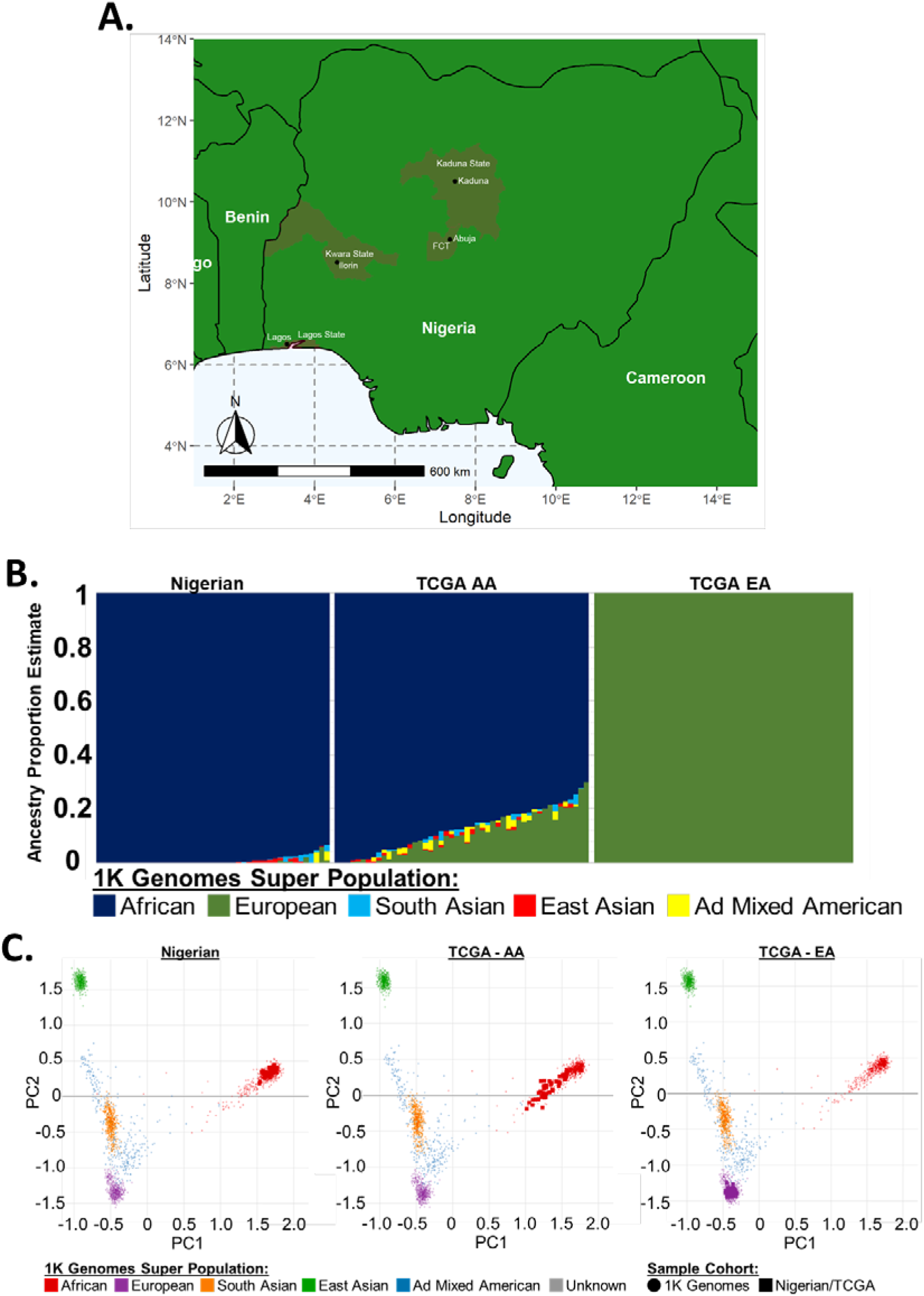
Sample Collection Sites and Genetic Admixture Analysis. **A)** At clinical sites across Nigeria, 45 samples were collected. **B)** Admixture v1.3.0 was used to estimate ancestry proportions, based on reference populations from the 1000 Genomes Project phase 3 superpopulations. Prior to analysis, rare variants (i.e., <5% across all phase 3 1000 genomes), all indels, and any SNPs that were not biallelic were removed. Samples within the CaPTC cohort had an average African proportion of 99.1%. TCGA Samples (n=50) with >70% African ancestry were classified as AA; 402 TCGA samples contained >60% European admixture. Those samples were sorted by European proportion, and the top 50 samples were classified as EA and utilized in this study. The average European proportion of this group was 99.996%. **C)** Germline variants within the NG and TCGA cohorts were compared to phase 3 1000 Genomes superpopulations using principal component analysis. NG samples strongly clustered with the African superpopulation. AAs samples clustered with the African superpopulation; European samples clustered with the European superpopulation.

### Pathological Scoring

The FFPE blocks were processed at the Pathology Biorepository Shared Service (PBSS) core at the University of Maryland Baltimore. For pathology review, an initial hematoxylin and eosin (H&E) slide was prepared from each block. A pathologist assessed the presence and quantity of tumor, and the presence and quantity of normal tissue. Up to two tumor and normal cores per H&E slide were circled, and the total number of cores per slide was recorded in a sample manifest. In addition, the pathologist provided the tumor core(s) Gleason score for each tumor circle. Those FFPE blocks with sufficient tumor and/or normal cores available were submitted to the PBSS research histology lab for core extraction. Using the corresponding H&E slide as a template, cores were extracted from each FFPE block by use of a manual tissue core extraction device with RNAse/DNAse-free conditions. The cores were placed in labelled RNAse/DNAse-free cryovials. A sample manifest for the extracted cores accompanied samples shipped to the NIH Laboratory of Human Carcinogenesis.

### Whole exome sequencing (WES)

As outlined in Supplemental Figure 1, library preparation was performed using Agilent SureSelectXT Human All Exon V6 r2 Exome Kits (Agilent Technologies, Santa Clara, CA, USA). Sequencing (2×150 bp) was performed on either an Illumina HiSeq4000 or on an Illumina NextSeq500 (Illumina, San Diego, CA, USA), to a target of 100 (± 10) million raw data reads per each sample library. Following sequencing, raw fastq files were transferred to the NIH Biowulf supercomputing cluster and analysed using the Center for Cancer Research Collaborative Bioinformatics Resource (CCBR) Whole Exome pipeline (https://github.com/CCBR/Pipeliner). Reads were trimmed using Trimmomatic v0.33^27^ and mapped to the hs37d5 version of the human reference genome (ftp://ftp.1000genomes.ebi.ac.uk/vol1/ftp/technical/reference/phase2_reference_assembly_sequence/hs37d5.fa.gz) using BWA-MEM v07.17^28^. Binary Alignment Map (BAM) files were processed using Samtools v1.8 (http://www.htslib.org/)^29^, and Picard v1 (http://broadinstitute.github.io/picard/) was used to mark duplicates. GATK v3.8^30^ was used to perform indel realignment and base recalibration. Read and alignment-level quality analysis was performed using Qualimap v2.2.1^31^. Alignment quality metrics were analysed and visualized using RStudio 1.2.5003 (http://www.rstudio.com) [R 3.6.3 (https://www.R-project.org)], the ggstatsplot v0.3.0 (https://indrajeetpatil.github.io/ggstatsplot/) package, and the compareGroups v4.2.0^32^ package.

### Variant Calling and Filtration

Germline variant calling was completed using HaplotypeCaller^33^, and Annovar v2019Oct24 was used for variant annotation. Variant filtration followed GATK Best practices. Cohort single nucleotide polymorphisms (SNPs) and indels were separated, filtered. and recombined for downstream analysis. SNP filters were Qual < 30.0, QD < 2.0, FS > 60.0, MQ < 40.0, MQRanksum < -12.5, and ReadPosRankSum < -8.0. Indel filters were Qual < 30.0, QD < 2.0, FS > 200.0, and ReadPosRankSum < -20.0. Once the variants were recombined, cohort germline variants were compared to a ClinVar-derived list of known PCa-associated germline variant regions (Supplemental Table 1) to target variants of known clinical importance. Germline variants with a read depth > 3 and a variant allele frequency > 50% were retained for downstream analysis. Somatic variant calling was completed using MuTect2^34^, and Annovar v2019Oct24^35^ was used for variant annotation. As described by Jones et al., a single unmatched NG normal FFPE exome, sequenced using the same methods as for the tumor samples, was paired with each tumor exome to filter false-positive variant calls^36^. Mutation tables were imported into RStudio for visualization and analysis using the maftools^37^ (v2.4.10) package. Variants were 1) screened for strand bias (Supplemental Figure 2) using GATK FisherStrand Phred Score, 2) separated into variants within known PCa-associated genes and variants within novel PCa-associated genes (Supplemental Table 2), using ClinVar and 3) and filtered (Supplemental Figure 3) using two filtering regimes. After filtering, retained variants within genes identified in ClinVar were considered “Known”; conversely, variants within genes not identified in ClinVar were called “Novel”. Filtering steps included 1) exclusion of silent and non-protein coding mutations, 2) variant allele read depth ≥3, 3) variant allele frequency >5% [10% for variants within novel PCa associated genes], 4) dbNSFP^38^ Genome Aggregation Database (gnomAD)^39^ exome allele frequency < 0.01 [< 0.001 for variants within novel PCa-associated genes and variants lacking allele frequencies were retained for downstream filtration], 5) identification as Pathogenic or Uncertain in ClinVar v 20200419^40^, 6) removal of dbSNP-annotated variants identified in NG unmatched normal samples (n = 11) (Supplemental Table 3), 7) present in genes altered in least 5% of tumors, and 8) manual validation in the Integrative Genomics Viewer (IGV) (Supplemental Figure 4). For genes mutated across at least five PCa samples, Fisher’s exact test was used to compare cohort mutation frequencies. The test was completed using the maftools clinicalenrichment function. p values < 0.05 were considered significant.

### TCGA PRAD Data Acquisition and Analysis

Access to The Cancer Genome Atlas (TCGA) PRAD data (Accession: phs000178.v11.p8) was obtained through the database of Genotypes and Phenotypes (dbGAP). Raw sequencing files in BAM format were downloaded through the Genomic Data Commons (GDC) data transfer tool from the GDC Data Portal (https://portal.gdc.cancer.gov/). After download, the raw files were sorted using Samtools and split into their constituent forward and reverse fastq files using bedtools v2.29^41^. Once separated, the fastq files were processed through the CCBR Whole Exome pipeline and analysed using the same methods as for NG CaPTC samples.

### Genetic Admixture Estimation

To ensure accurate ancestral group assignment, HaplotypeCaller^42^ and Admixture v1.3.0^43^ were used to estimate ancestry proportions, based on reference populations from the 1000 Genomes Project phase 3 superpopulations in all TCGA and NG samples (Supplemental Figure 5). Rare variants (i.e., <5% across all phase 3^44^ 1000 genomes), all indels, and any SNPs that were not biallelic were removed prior to analysis. TCGA samples (n=57) with majority African ancestry were classified as AAs. Only the samples with >70% African ancestry (n=50) were retained for comparison to NG samples. 402 TCGA samples contained >60% European admixture. Those samples were sorted by European proportion, and the top 50 samples were classified as EAs and utilized in this study. The average European proportion of this group was 99.996%.

### COSMIC Signature Enrichment

Using the maftools package, filtered single-nucleotide variants across each cohort were used to estimate the representation of Catalogue of Somatic Mutations in Cancer (COSMIC) (cancer.sanger.ac.uk)^45^ mutation signatures within each tumor sample. Maftools uses cophenetic correlation and nonnegative matrix factorization to determine the optimal number of SNP signatures (across the cohort), extracts those signatures, and compares them to the known (n=30) COSMIC signatures.

### Variant Functional Gene Ontology and Network Analyses

Filtered variants, present in at least two NG PCa tumor samples, were imported into Cytoscape^46^ (v. 3.7.2) to assess functional gene ontology enrichment and to visualize the GO term interaction network. Once separated, functional analysis and network construction were completed using the stringApp (v 1.6.0)^47^ and ClueGO (v. 2.5.7) plug-ins^48^. A two-sided (enrichment/depletion) hypergeometric test with Bonferroni step-down was used to determine Reactome Pathways (v. 08.05.2020) enrichment. Analysis thresholds included enrichment significance of p ≤ 0.01, a minimum of 5% gene inclusion and a kappa score threshold of ≥ 0.4. ClueGO uses kappa scores to determine the likelihood of GO term interactions and groupings.

## Results

To determine the mutations in PCa of NG men collected from multiple institutions within the CaPTC, we focused on 45 intermediate (Gleason scores 4+3) and high-grade (Gleason scores ≥ 4+3) tumors obtained across multiple institutions (the NG cohort was >68X, with an average mapping quality of 57.2 and an average mapped-read count of 263 million per sample) (Supplemental Table 4). 20 samples were collected from Northern Nigeria), 17 samples from Central Nigeria, and 8 samples from Southwest Nigeria (Supplemental Table 5). PRAD exome data were downloaded from TCGA, using the dbGAP database as a comparison cohort (Supplemental Table 6).

Race is a poor group classifier for linking genetic variation and disease causation^49^; moreover, self-reported race can obscure genetic variation due to misunderstandings about family heritage, cultural influences, and/or other societal factors^50^. To ensure that our NG and TCGA cohort comparisons were not skewed by bias within self-reported race, we quantified the individual genetic admixture within each patient sample. To accomplish this, germline SNPs were compared to 1000 genomes super populations (African, European, South Asian, East Asian, and admixed American), and ancestry proportion estimates were calculated (Figure 1B). NG patients showed an average genetic ancestry of 99.1% African. The genetic ancestry of TCGA AA patients was predominantly a mixture of African (50.2%-99.99%) and European admixture (1%-43%). To reduce the impact of this variance on our analysis, we selected only TCGA AA patients with ≥70% African ancestry (n=50). TCGA EA patients showed minimal admixture, with >98.3% European ancestry. Nine patients self-identified as EAs possessed ≤45% European ancestry. Four of the nine patients were majority (>50%) admixed American, two were majority East Asian, two were majority African, and one was predominantly (45%) European with 35% admixed American and 16% African admixture. To obtain an EA comparison group, we sorted (high to low) the cohort by European ancestry proportion and selected the top 50 TCGA EA patients. After admixture estimation and sample selection, principal component analysis plots were used to visualize the relationships between each cohort and the five 1000 Genomes superpopulations (Figure 1C). The NG and TCGA EA cohorts clustered with their ancestral 1000 genomes superpopulations, and the TCGA AA cohort clustered with the African superpopulation. Thus, data for these patients were used in subsequent analyses.

The NG cohort harboured 31 known, non-benign, germline variants. Four genes [*BRCA1* (100%), *BARD1* (45%), *BRCA2* (27%) and *PMS2* (18%)] were altered in at least two samples (Figure 2A). These genes also showed top mutation frequencies within both TCGA cohorts (Figure 2B and C). Across 111 germline variants, the AA TCGA cohort reflected a pattern [*BRCA1* (68%), *BARD1* (34%), *BRCA2* (28%), and *PMS2* (16%)] similar to that for NG samples. Additionally, the rate of *BRCA1* alterations increased (p ≤ 0.021) as African admixture increased (Supplemental Figure 6). 126 germline variants were present in the EA TCGA cohort. Disaggregating mutation frequencies down to specific variants revealed both ancestry-linked and NG-specific germline variant patterns. *BRCA1* showed an increasing mutation frequency as African admixture increased (Figure 3A). That pattern was driven by three variants [rs799917, rs16941, and rs16942] (Figure 3B). The frequency of rs799917 was higher for men of African ancestry; rs16941 and rs16942 were lower. In esophageal squamous cell carcinoma, the BRCA1_rs799917 T>C SNP inhibits mir-638-mediated regulation of BRCA1, thus reducing BRCA1 expression and increasing cancer cell proliferation^51^. This variant is also linked to a higher risk of gastric, lung, and triple-negative breast cancer^52-54^. BRCA1_rs16941 and BRCA1_rs16942 are variants of unknown significance (VUS). *BARD1* germline variant patterns appear to be specific to NG men. Compared to AA and EA cohorts, rs2070096 is lower, and rs2070094 is higher. Of note, the BARD1_rs2070094 SNP resides within the BARD1-binding domain of *BRCA1* and may provide a protective function that enhances DNA repair by enhancing BARD1/BRCA1 binding stability^55^. BARD1_rs2070096 is a VUS. *BRCA2* germline variants displayed both ancestry-linked and NG-specific patterns. rs11571831 was present only in men of African ancestry, and rs766173 was high in NG men. Both *BRCA2* variants are classified as VUS. Although most variants in NG PCa were identified as VUS, their presence and differing frequencies, compared to TCGA, provide opportunities for future investigations. Characterizing the full mutational spectrum is a first step in understanding how best to diagnose and treat this underrepresented patient population.

**Figure 2.**
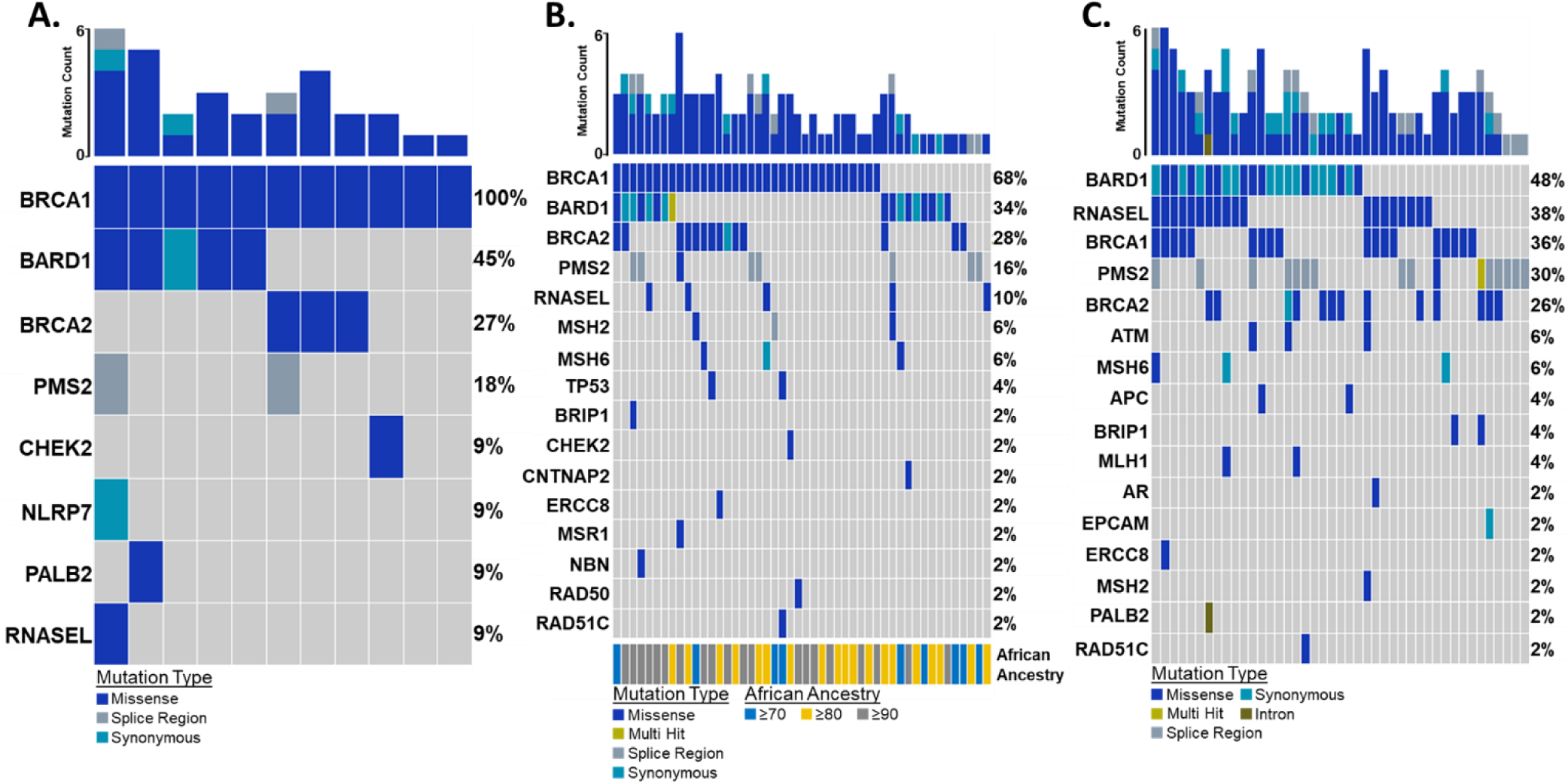
PCa Germline Variant Oncoplot. **A)** NG germline variants detected across 11 normal samples were filtered against known ClinVar cancer variants. In at least two tumor samples, four genes known to harbor cancer-related variants were mutated. These genes included *BRCA1* (BRCA1 DNA Repair Associated) – 100%, *BARD1* – 45%, *BRCA2* (DNA Repair Associated) – 27%, and *PMS2* (PMS1 Homolog 2, Mismatch Repair System Component) – 18%. As a comparison to NG PCa exome samples, TCGA PCa samples (n = 50 AA and n = 50 EA) were downloaded through dbGAP and analyzed for genetic variants. **B)** In the AA cohort. eight genes showed germline mutations in at least two samples. **C)** In the EA cohort, ten genes showed germline mutations in at least two samples.

**Figure 3.**
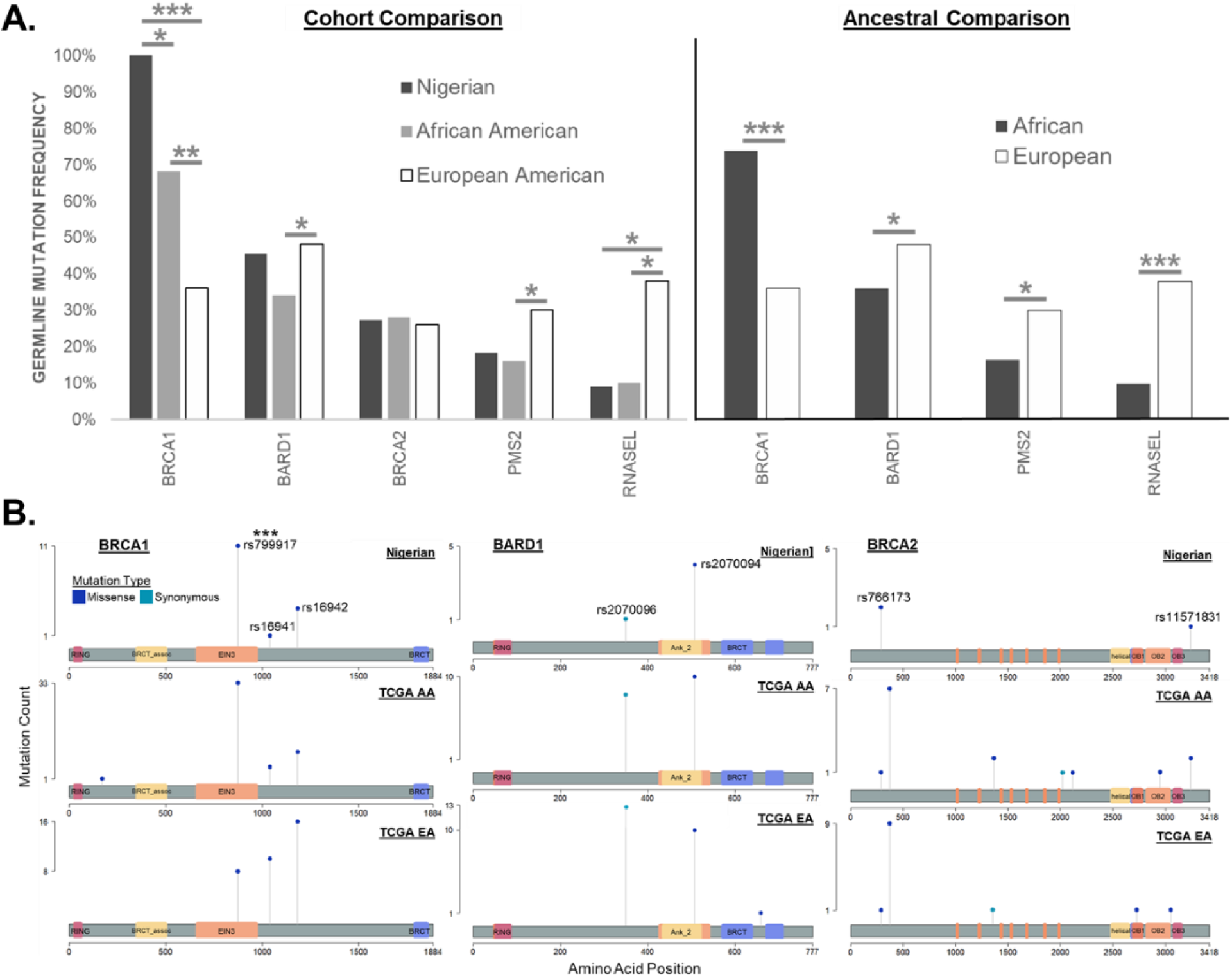
NG PCa Cohort Germline Mutations Comparison to TCGA PCa Cohorts and Lollipop Plots. **A)** PCa of NG and AA men showed more *BRCA1* germline mutations (p ≤ 0.001 and p ≤ 0.01, respectively) compared to European men. Additionally, PCa of NG men showed more (p ≤ 0.036) BRCA1 mutations relative to AA men. In PCa of EA men, *BARD1* was mutated at a higher rate (p ≤ 0.048). *BRCA2* showed no significant difference in cohort mutation rates. **B)** To disaggregate mutation rates down to specific variants, lollipop plots revealed a finer variation in cohort patterns. The ancestry-linked pattern of *BRCA1* is driven by rs799917, which was more frequent (p ≤ 0.001) for men of African ancestry. rs16941 and rs16942 were elevated in PCa of European men, but that difference was not statistically significant. *BARD1* germline variants showed no significant difference in variant rates; however, the patterns appeared to be specific to NG men. Compared to AA and EA cohorts, rs2070096 was lower, and rs2070094 was higher. *BRCA2* germline variants displayed no statistically different variant rates, but both ancestry-linked and NG-specific patterns were discernable. rs11571831 was present only in PCa of men of African ancestry, and rs766173 was elevated in PCa of NG Men. P values were produced via two-sided Fisher’s exact test groupwise comparison.

Somatic variant analysis of tumor-only sequencing data involves multiple nontrivial steps that are distinct from the analysis of matched tumor and normal sequencing. Therefore, we used an established pipeline that incorporated a panel of normal samples. We used an unmatched NG normal sample to filter out NG-specific germline variants^36,56^, reducing the unique NG variants by 70.8% from 2,506,254 to 730,285 variants (Supplemental Figure 7). Within the TCGA cohorts, we used each sample’s patient-matched normal, which produced 11,208 unique AA variants and 15,191 unique EA variants. Since the NG cohort contained many somatic variants, we employed two filtering regimes, one for variants within known PCa-associated genes (as identified in ClinVar) and one for variants within genes not associated with PCa. We identified 905 variants across 25 genes known to be associated with PCa, and 156 variants across 51 novel PCa genes. Using the same approach, we identified 15,854 variants in the TCGA AA cohort and 21,957 variants in the TCGA EA cohort. Consistent with other sequencing studies^57^, our results showed the same racial mutation patterns for *SPOP, ATM, TP53*, and *PIK3CA*. TCGA cohorts did not show recurrent mutations in genes not associated with PCa. Our dual filtering approach allowed us to filter, independently, each set of variants across the NG cohort without over-filtering variants within known PCa-associated genes and to identify high-confidence variants in novel PCa-associated genes.

Within the NG cohort, 133 somatic variants were present in 26 PCa-associated genes. Nine genes [*BRCA2* (27%), *APC* (20%), *ATM* (20%), *BRCA1* (13%), *DNAJC6* (13%), *EGFR* (13%), *MAD1L1* (13%), *MLH1* (11%), and *PMS2* (11%)] showed mutation frequencies > 10% (Figure 4A). Of NG PCa, 53% showed mutations in genes (*BRCA2, ATM, BRCA1, CHEK2, TP53*, and *MSH6*) associated with genome integrity. The TCGA AA and EA cohorts harbored 67 and 73 somatic variants, respectively. Across both cohorts, fifteen genes were mutated in at least 2 samples [*SPOP, ATM, TP53, BRAF, MED12, PIK3CA, CTNNB1, EGFR, FLCN, MYH7, PTEN*, and *TTN*]. *SPOP* and *ATM* were the most frequently mutated genes in AA tumors and were mutated two times more compared to EA. Comparison of the mutation frequencies between TCGA cohorts did not show any statistically significant differences; however, AA tumors showed a significant increase in *SPOP* mutations compared to NG (Figure 4B). *BRCA2, APC*, and *BRCA1* showed statistically significant increases in the NG cohort. Though not statistically significant, *ATM* had the highest mutation frequency associated with increasing African ancestry; specifically TCGA EAs had an *ATM* mutation frequency of 4%, but TCGA AA and NG cohorts had rates of 8% and 20%, respectively. Somatic mutations for NGs and AAs were distributed across the amino acid sequence of the most mutated genes. None of the variants were shared across or within cohorts. (Figure 4C).

**Figure 4.**
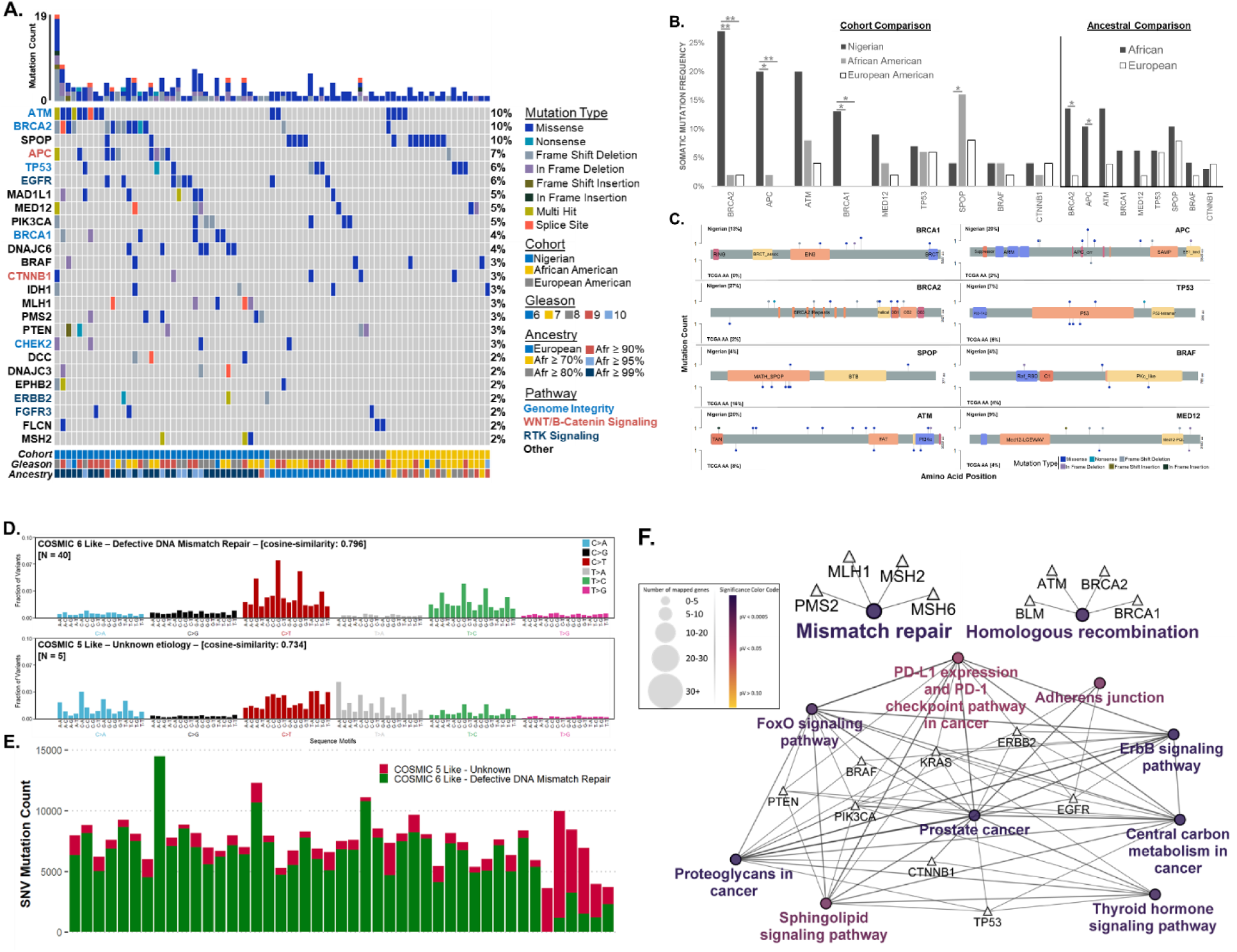
PCa Somatic Variants within Known PCa-Associated Genes. **A)** Variant calling within the NG cohort (n=45) produced 1,168,250 variants. 25 genes were known to be associated with PCa harbored variants in at least two tumor samples. The most frequently mutated of these included *BRCA2* (BRCA2 DNA Repair Associated) – 27%, *APC* (APC Regulator of WNT Signaling Pathway) – 20%, *ATM* (ATM Serine/Threonine Kinase) – 20%, *BRCA1* (BRCA1 DNA Repair Associated) – 13%, and *DNAJC6* (DnaJ Heat Shock Protein Family (Hsp40) Member C6) – 13%. As a comparison to NG PCa exome samples, TCGA PCa samples (n = 50 AA and n = 50 EA) were downloaded through dbGAP and analyzed for genetic variants. **B)** PCa of NG men showed a significant (p ≤ 0.01) elevation in *BRCA2* somatic mutations compared to African and EA men. A significant increase was also evident for *APC* (p ≤ 0.05) and *BRCA1* (p ≤ 0.05). Compared to NG men, PCa of AAs were elevated (p ≤ 0.05). NG and AA men also showed higher, but not significant, mutation frequencies of *ATM, MED12*, and *BRAF*. **C)** Somatic mutations for NGs and AAs were distributed across the amino acid sequence of the most mutated genes. None of the variants were shared across or within cohorts. **D and E)** SNPs in the NG PCa cohort were compared to known cancer-related mutation signatures within the Catalogue of Somatic Mutations in Cancer (COSMIC). 89% of NG PCa mutation patterns were similar (cosign similarity ≥ 0.796) with COSMIC signatures 6. The remaining 11% were more like COSMIC 5. **F)** Mutated genes (n=83) present in at least two NG PCa tumor samples (n=45) were imported into Cytoscape to assess functional gene ontology enrichment and visualize the GO term interaction network, using KEGG pathways. Variants showed significant (q ≤ 0.000538) GO and functional enrichment across multiple GO groups, including mismatch repair, homologous recombination, PCa, and several cancer-related signaling pathways.

In addition to the variants within known PCa driver genes, we identified four novel mutated genes that showed mutation frequencies > 10%. *CACNA2D2* had the highest mutation rate of 29% (Supplemental Figure 8A) and showed a recurrent (n=13) missense SNP of Leu54Phe (rs569543350) (Supplemental Figure 8B). *TTN* (Titin) had the second highest mutation frequency at 20%. The size of this large protein (>30000 amino acids) renders it more susceptible to DNA repair errors, making the functional significance of these mutations unreliable, even after rigorous variant filtering^58-61^. *SYNE1* was the third most frequently mutated gene (16%) in the novel PCa set. This gene showed a recurrent (n=2) missense SNP of Gln1491Glu. The fourth most mutated gene was *ADAMTS2*. This gene showed a recurrent (n=4) in-frame insertion of Leu_Pro29dup and an overall mutation frequency of 13%. Lastly, 47 other genes not known to be associated with PCa were mutated in two or more patients; however, we did not characterize these due to their low mutation frequencies.

To validate the observed germline variant frequencies, we analyzed NG tumors using the germline variant pipeline (Supplemental Figure 11). NG tumor samples not only possessed comparable rates of gene level mutation [*BRCA1* (100%), *BARD1* (41%) and *BRCA2* (18%)] but also showed comparable variant frequencies.

We next investigated the overall mutational patterns within each cohort to understand global somatic events. The NG cohort shared similarities (cosign similarities ≥ 0.734) with COSMIC signatures 5 and 6 (Figure 4D). Five NG cohort samples had a mutational pattern similar (cosign similarity ≥ 0.734) to COSMIC 5. Forty cohort samples were similar (cosign similarity ≥ 0.796) to COSMIC 6. TCGA AA mutational patterns shared similarities with COSMIC 1 and 5 (cosign similarities ≥ 0.645), and TCGA EA tumors shared similarities with COSMIC 1, 3, and 4 (cosign similarities ≥ 0.481) (Supplemental Figure 10). Within the COSMIC database, mutational signatures 1, 5, and 6 were the signatures most often observed in PCa.

To determine the mechanism associated with tumorigenesis in NG PCa tumors, mutated genes present in at least two NG patients were analysed for functional gene ontology enrichment. NG tumors showed significant (q < 0.001) GO and functional enrichment in mismatch repair and non-homologous repair deficiency pathways (Figure 4F and Supplemental Table 7). Additional enriched pathways included PD-1 checkpoint, thyroid hormone signalling, FOXO signalling, ErB2 signalling, adherens junctions, proteoglycans, and sphingolipids.

## Discussion

This study is the first to perform whole-exome sequencing of advanced-stage, treatment-naïve primary tumors from NG men with PCa. We analyzed the genomes of 45 tumors and compared them to a reference panel of normal samples. Since most AAs in the US have West African Ancestry, we assessed genetic admixture on comparison data sets from TCGA, which contains data on AA and EA PCa patients. We evaluated ancestry-linked germline and somatic alteration frequencies in DNA damage-repair genes (*BRCA1, BRCA2, APC*, and *ATM*), as well as three novel PCa-associated genes (*CACNA2D2, SYNE1*, and *ADAMTS2*). Alterations in DNA damage repair pathways are involved in PCa development and progression and are clinically targetable^62^. Since men of African ancestry are severely under-represented in genomic studies, our findings address a gap in the contribution of genetic variation to the incidence of PCa and aggressiveness of the disease in MAAs. Further, these findings encourage us to identify clinically targetable sites to close the gap in health-related disparities.

We observed a high level of BRCA1 germline mutations in PCa of NG and men of African ancestry. The high rate is driven by the variant BRCA1_rs799917 T>C, which enhances disease risk in triple-negative breast^54^, gastric^52^, esophageal squamous cell^51^, and lung^53^ cancers. Results of meta-analyses, however, suggest that this variant is non-pathogenic^63-65^. Since none of these studies included patients of African ancestry, the impact of this variant on that population remains poorly explored. BRCA1_rs799917 T>C alters the coding sequence of *BRCA1*, lowering BRCA1 expression by inhibiting its interaction with miR-638^66^. Lower expression of *BRCA1*, a DNA damage repair gene, leads to accumulation of mutated DNA, which enhances tumorigenesis. BRCA1_rs799917 T>C has not previously been associated with PCa; however, *BRCA1* germline mutations contribute to increased PCa risk^67^ and are associated with higher PCa aggression and poorer outcomes^68^. We found this trend in AA PCa, which have higher frequencies of germline *BRCA1* VUS^69^. This pattern is evident in both NG and AA breast cancer tumors^70-72^. Thus, our exome analysis provides evidence of distinctive germline *BRCA1* alterations in PCa of patients with African Ancestry.

Our analysis also showed that, in their DDR genes, NG PCa tumors have higher somatic mutation rates, with 53% of NG tumors having at least one somatic DDR gene mutation. NG tumors demonstrated increases in *BRCA2, APC*, and *BRCA1* mutations and SNP patterns associated with defective DNA mismatch repair. In addition, these tumors contained mutated genes that had significant gene ontology and functional enrichment across multiple GO groups, including mismatch repair and homologous recombination signalling pathways. Pathogenic alterations in DDR genes are prevalent in advanced-stage, localized PCa, especially affecting genes responsible for repair by homologous recombination (HR)^73^. Of note, BRCA2 facilitates the formation of RAD51 (RAD51 Recombinase) filaments, which are necessary for HR^74^. The clinical implications of somatic *BRCA2* mutations are poorly understood; however, all *BRCA2* mutations (germline and somatic) are understood to destabilize HR, increase tumor aggression, and contribute to poor patient outcomes^75^. DNA repair signalling is often impaired in cancer cells^76^; however, this impairment is more substantial for MAA. Yadav et al. observed increased somatic mutations of *BRCA2, BRCA1*, and *ATM* in AA PCa^24^. AAs had a 1.24-to 2.16-fold increase in *BRCA2, BRCA1*, and *ATM*, as compared to EAs. The innate impairment of DNA repair in cancer cells leads to a dependence on alternative repair pathways that can be therapeutically exploited. Taking our findings, which are in line with other reports for AA men, the recent FDA approval of PARP inhibitors (specifically olaparib) and other therapies such as platinum drugs, ATR inhibitors, CHEK1/2 inhibitors, and radiotherapy may be useful for men of African Ancestry^77,78^.

Our analyses identified, in addition to somatic alterations in DDR genes, somatic mutations in novel PCa-associated genes. Of NG tumors, 27% (n=13) contained a recurrent *CACNA2D2* missense SNP of Leu54Phe (rs569543350). Both ClinVar and dbSNP designate this variant as having “unknown significance”. *CACNA2D2* modulates the expression of functional calcium channels^79^, which contribute to cancer development^80^. Compared to non-cancerous prostate tissue, *CACNA2D2* is expressed higher in PCa tissue and can increase tumor proliferation and angiogenesis^81^. *SYNE1* encodes a multi-isomeric protein that participates in connecting the nuclear envelope to the cytoskeleton. This connection is necessary for proper nuclear movement and positioning, and for cellular migration^82^. Abnormal nuclear envelope structure, a feature of cancer, is thought to contribute to tumorigenesis^83^. Mutations in *SYNE1* are linked to several human cancers^84^. *ADAMTS2* encodes a procollagen N-proteinase that is necessary for collagen fibril assembly^85^. The role of *ADAMTS2* in cancer remains poorly understood; however, the impact of collagen metabolism is well-characterized^86,87^. Collagen is a structural component of the extracellular matrix, and its metabolism can affect tumor development, tumor tissue stiffness, metastasis, and treatment response. The presence of these novel PCa-associated alterations is unsurprising. Non-European populations have higher rates of VUS^88-90^. This is a result of the lack of diversity in research and (by extension) genomic databases^91^. Non-European populations are underrepresented in research and genomic databases, which skews genomic annotations away from identifying clinically relevant variants in these populations. Further investigation into these novel alterations may expose alternative routes of disease aggression for MAA and mitigate the disparities.

These findings present the most comprehensive characterization of the NG PCa exome to date, and highlight the need to increase study population diversity. Although clinical genomics is a powerful tool to guide clinical interventions, the lack of non-European patients limits the capacity of these advancements to benefit men of African ancestry. Furthermore, the high level of genetic diversity within African men necessities the need for larger cohort studies to identify population-specific, recurrent mutations that contribute to prostate carcinogenesis.

Although our results are compelling, the study has some limitations. The high level of genetic diversity in Africans coupled with the lack of matched normal samples increased the number of detected somatic variants, requiring us to aggressively variant filter variants to limit the possibility of false positives. This conservative approach, although necessary, provides the possibility that we inadvertently excluded some true somatic variants. In addition, the limited representation of African patients within genomic databases and genomic research reduces our ability to determine the larger population-specific distribution of these findings. To our knowledge, this is the largest PCa exome study of NG men, and, despite the limitations, provides a robust characterization of the somatic landscape within NG PCa.

## Data Availability

All data produced in the present study are available upon reasonable request to the authors. Data deposition into dbGap is in progress.

## Author Contributions

CY, FTO, KAA, PJ, JOO, CNO, MF, AP, OAF and OPO conceived and designed the study and collaborated to collect the NG FFPE samples. DYG assisted with sample collection. ETK organized sample manifests, managed sample data and compiled preliminary sequencing results. DKF assisted with sample data management. BK coordinated sample preparation for pathology. DNM provided funding to complete WES of the collected samples. KRB coordinated sequencing design, completion, and data deposition. SA provided resources for data analysis, dbGap access and methodological consultation. JW organized data for analysis, ran analysis, and wrote the manuscript in conjunction with CY and SR. IE, WT, MD, PP and MJC contributed to data analysis and interpretation. All authors revised and approved the manuscript.

## Acknowledgements

We thank all members of the Prostate Cancer Translatlantic Consoritium (including our community partners) and members of each respective lab that contributed to this study. We also thank members of the CCR Collaborative Bioinformatics Resource group (Mayank Tandon and Skyler Kuhn), Justin Lack of the NIAID Collaborative Bioinformatics Resource group, and numerous staff within the NIH High-Performance Computing group.

## Supplemental Figure Legends

**Supplemental Figure 1.**
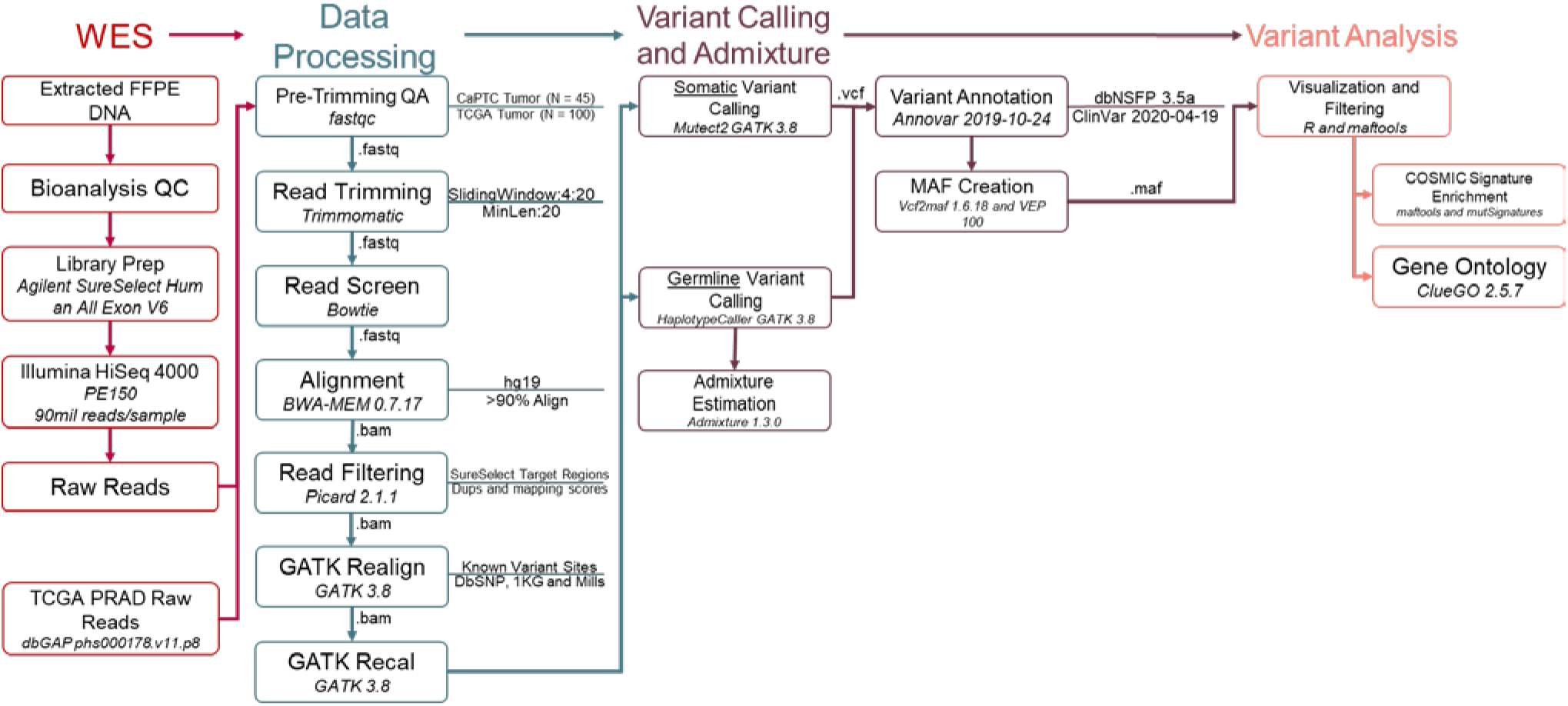
WES Analysis Workflow. NG CaPTC and TCGA PCa DNA samples were processed and analyzed using the CCBR WES pipeline in tumor-normal somatic variant and germline modes. Incorporating GATK Best Practices, CCBR expertise, and project specific modifications, these pipelines provided comprehensive raw data quality assurance, variant calling, and genetic admixture estimation. Output mutation tables (maf) and variant calls (vcf) were used for all downstream analyses. CaPTC cohort tumor exomes were paired with a single unmatched normal exome to diminish false-positive somatic variant calls. Patient-matched normals were used for TCGA exomes.

**Supplemental Figure 2.**
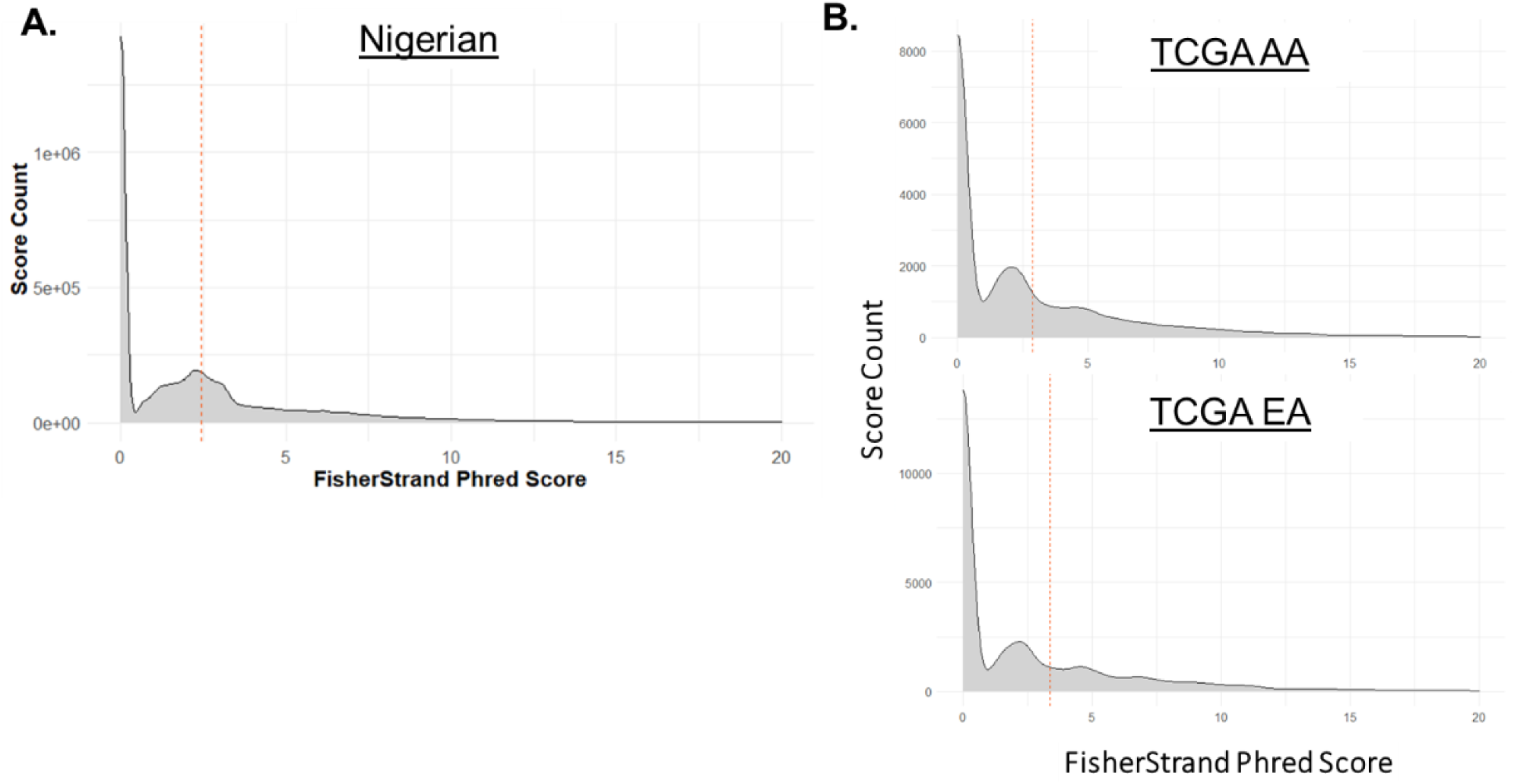
Variant Strand Bias. Variant annotation included the FisherStrand metric generated by Mutect2. This phred-scaled Fisher’s Exact Test p-value provides an indicator of strand bias probability. GATK best practices uses an FS score of ≤60 to filter out variants with a high probability of strand bias. Within the NG cohort **A)**, FS scores were well below that threshold, meaning the probability of the reads being false positive is diminished. FS scores within AA **B)** and EA **C)** TCGA samples were also below that threshold. The red dashed line denotes the mean FisherStrand Phred score.

**Supplemental Figure 3.**
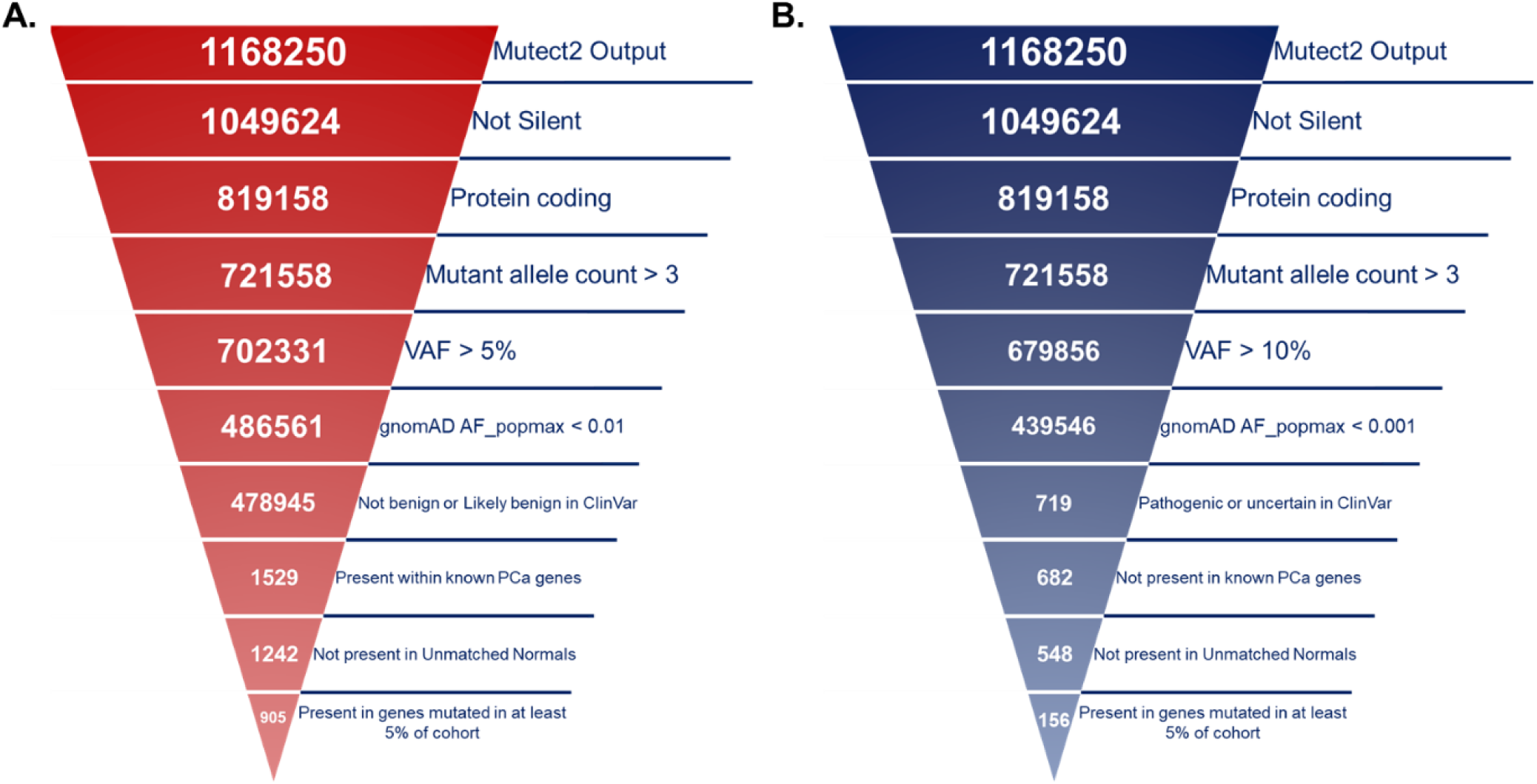
Somatic Variant Filtering. Two filtering regimes were utilized to identify NG PCa somatic variants within known and novel PCa-associated genes. **A)** To identify variants within known PCa-associated genes, mutated NG cohort genes not identified as harboring pathogenic somatic mutations in ClinVar were excluded, with remaining variants being filtered conservatively. **B)** Variants within novel PCa-associated genes were aggressively filtered and were not in genes identified in ClinVar.

**Supplemental Figure 4.**
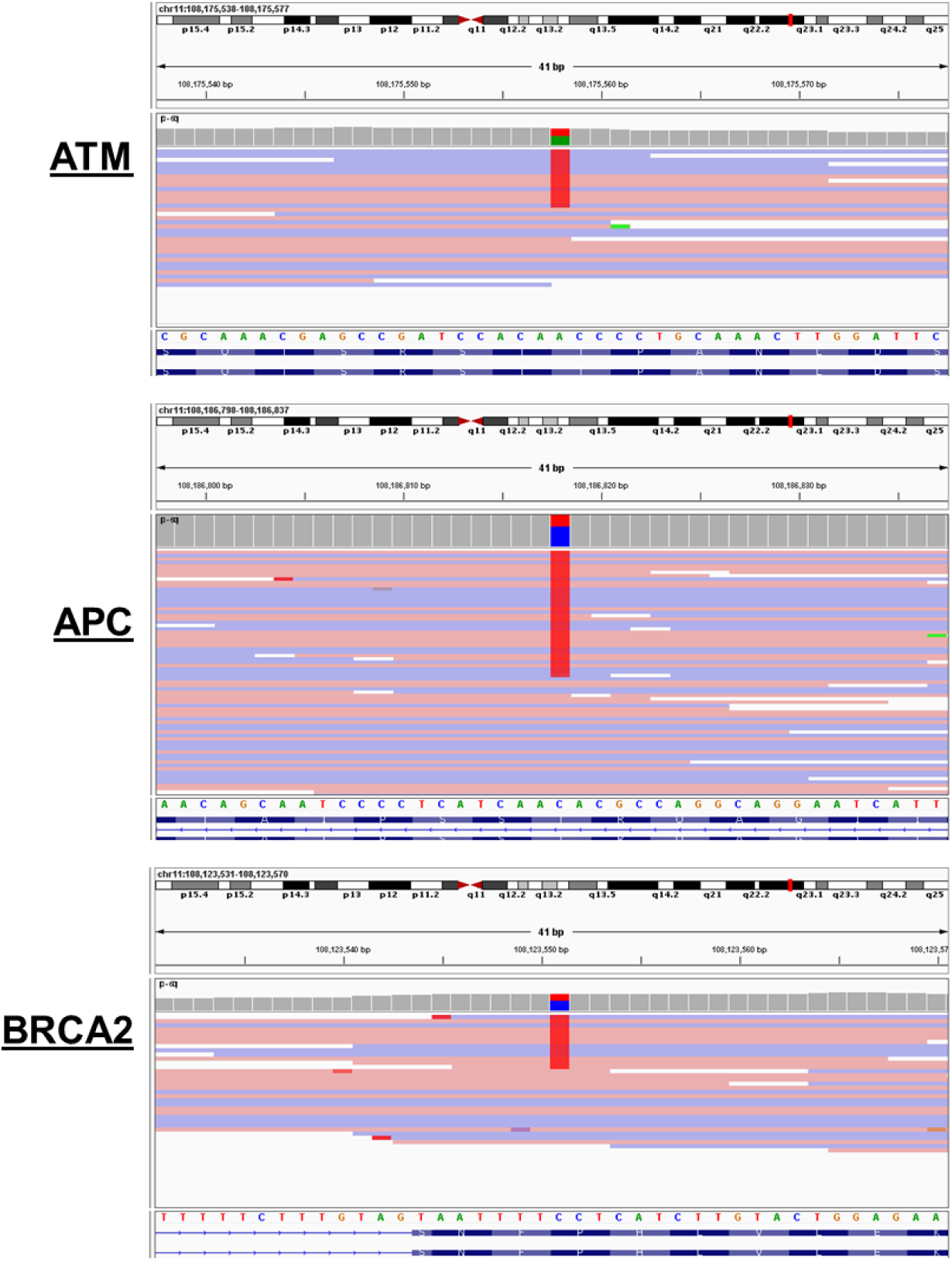
Manual Somatic Variant Inspection. Following variant filtration, variants within the most frequently mutated genes were manually inspected in IGV to validate variant calls. If a nucleotide differs from the reference sequence in greater than 20% of quality weighted reads, IGV colors the bar in proportion to the read count of each base (A, C, G, T). A, green; C, blue; G, yellow; T, red.

**Supplemental Figure 5.**
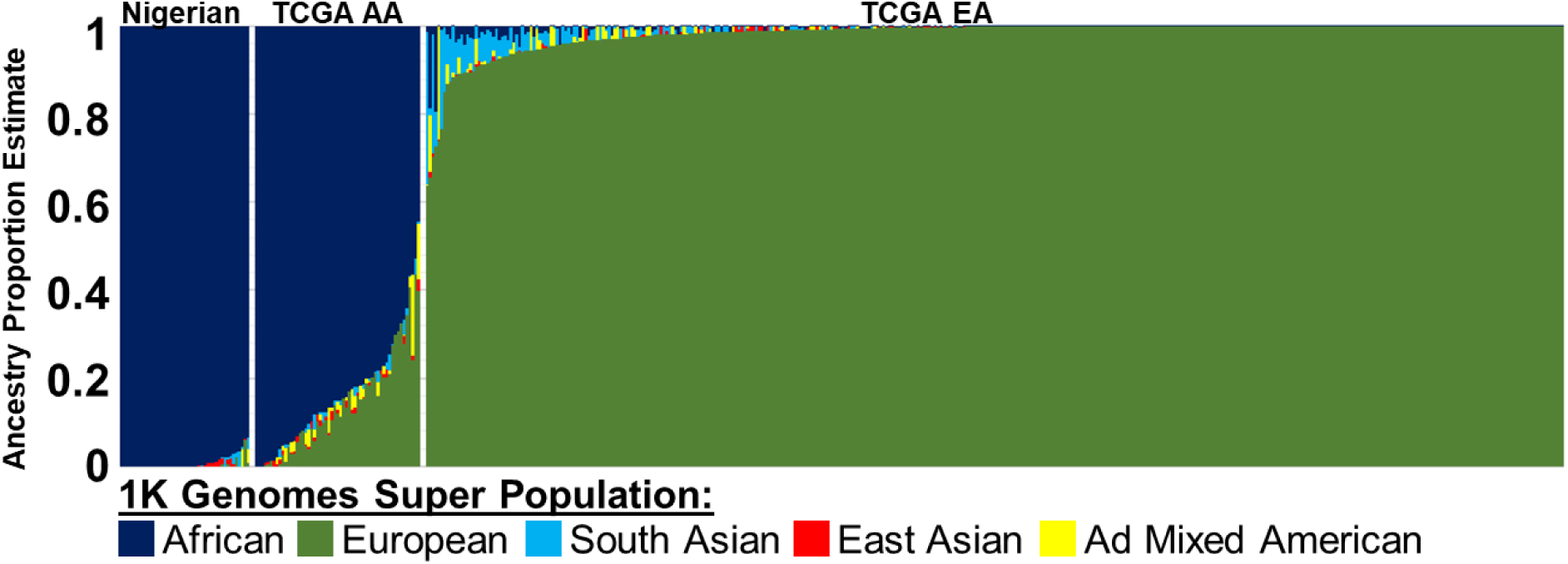
Genetic Admixture Analysis. Admixture v1.3.0 was used to estimate ancestry proportions, based on reference populations from the 1000 Genomes Project phase 3 superpopulations. Rare variants (i.e., <5% across all phase 3 1000 genomes), all indels, and any SNPs that were not biallelic, were removed prior to analysis. Samples within the CaPTC cohort had an average African proportion of 99.1%. TCGA Samples (n=50) with >70% African ancestry were classified as AAs. 402 TCGA samples contained >60% European admixture.

**Supplemental Figure 6.**
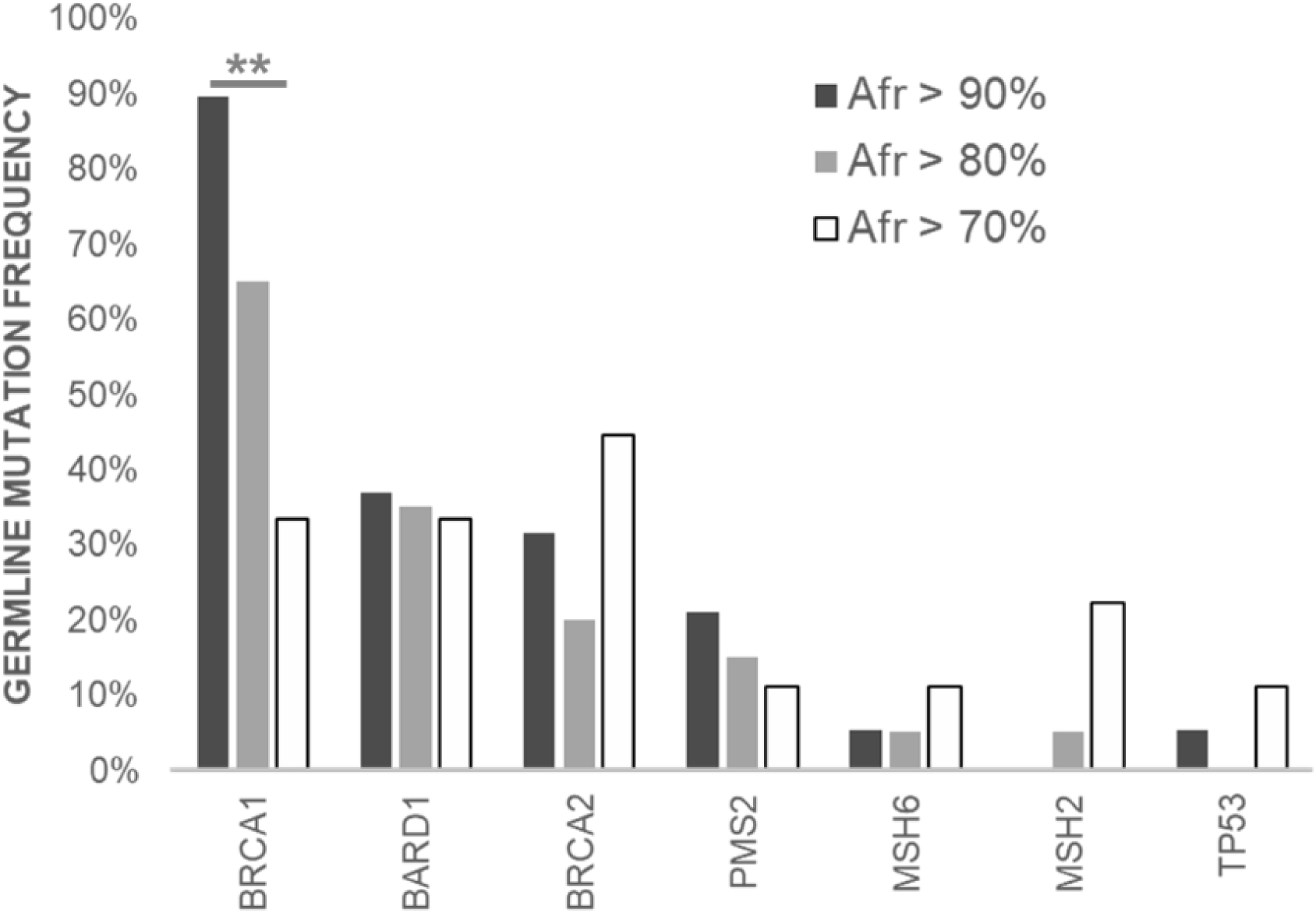
AA PCa TCGA Cohort Germline Mutation Comparison by African Ancestry. AA men with > 90% African ancestry (n = 19) showed a significantly higher (p ≤ 0.021) number of *BRCA1* germline mutations relative to those with lower amounts of African ancestry (n = 29). Additionally, AA men with >90% African ancestry had a statistically higher (p = 0.012) frequency of the *BRCA1* variant rs799917.

**Supplemental Figure 7.**
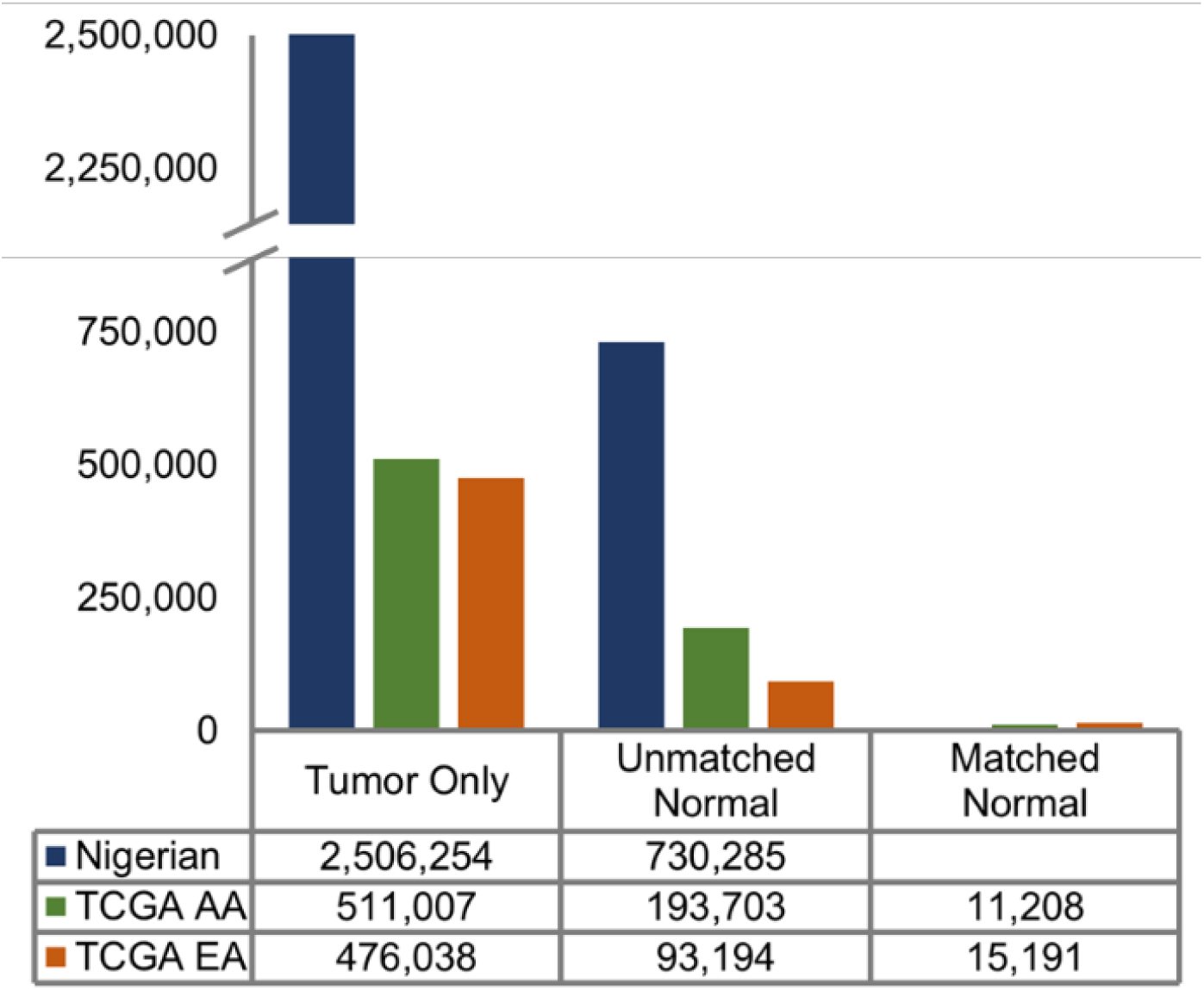
Comparison of somatic variant count based on normal sample usage. To determine the impact of normal sample usage on the total number of called variants, each cohort was analyzed up to three times. NG CaPTC samples were analyzed using two methods (1-Tumor-only and 2-Against an unmatched normal). Tumor-only analysis produced 2,506,254 variants; however, that count was reduced by 70.8% when an unmatched normal was used. Each TCGA cohort was analyzed using three methods (1-Tumor only, 2-Against an unmatched normal, and 3-Against a matched normal). For AA TCGA samples, tumor-only analysis produced 511,007 variants; however, that count was reduced by 62.1% and 97.8% with unmatched and matched normals, respectively. For EA TCGA samples, tumor-only analysis produced 476,038 variants; however, that count was reduced by 80.4% and 96.8% with unmatched and matched normals, respectively.

**Supplemental Figure 8.**
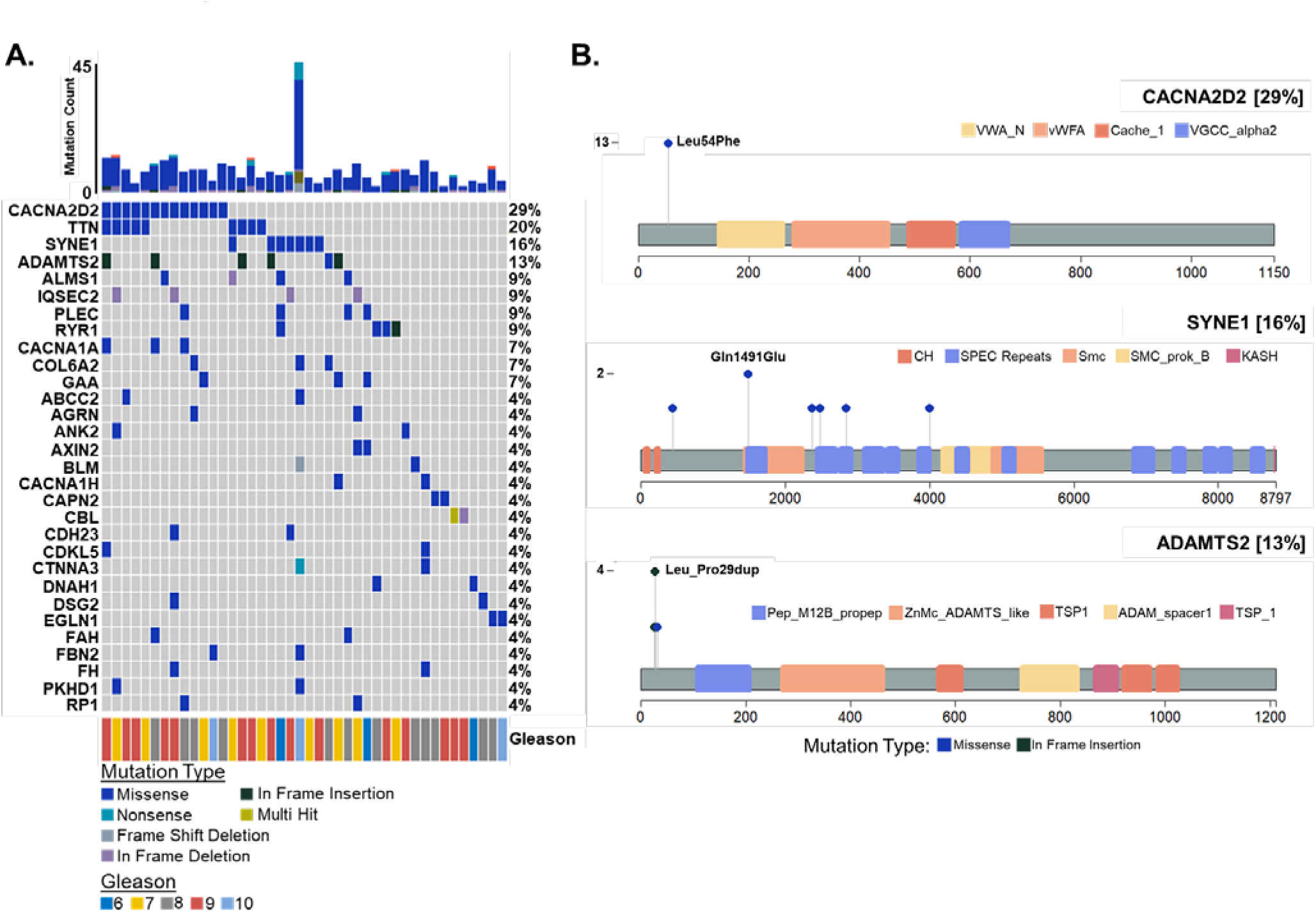
NG PCa Novel Somatic Variants. Variant calling within the NG cohort (n=45) produced 1,168,250 variants. NG PCa variants not within canonical genes, identified in ClinVar, were retained and stringently filtered. **A)** 51 genes with novel PCa association harbored variants in at least two tumor samples. **B)** *CACNA2D2* (Calcium Voltage-Gated Channel Auxiliary Subunit Alpha2delta 2) had the highest cohort mutation rate of 29%, showing a recurrent (n=13) missense SNP of Leu54Phe. *SYNE1* (Spectrin Repeat Containing Nuclear Envelope Protein 1) had a mutation frequency of 16% and a recurrent (n=2) missense SNP of Gln1491Glu. *ADAMTS2* (ADAM Metallopeptidase with Thrombospondin Type 1 Motif 2) showed a recurrent (n=4) in-frame insertion of Leu_Pro29dup and an overall mutation frequency of 13%.

**Supplemental Figure 9.**
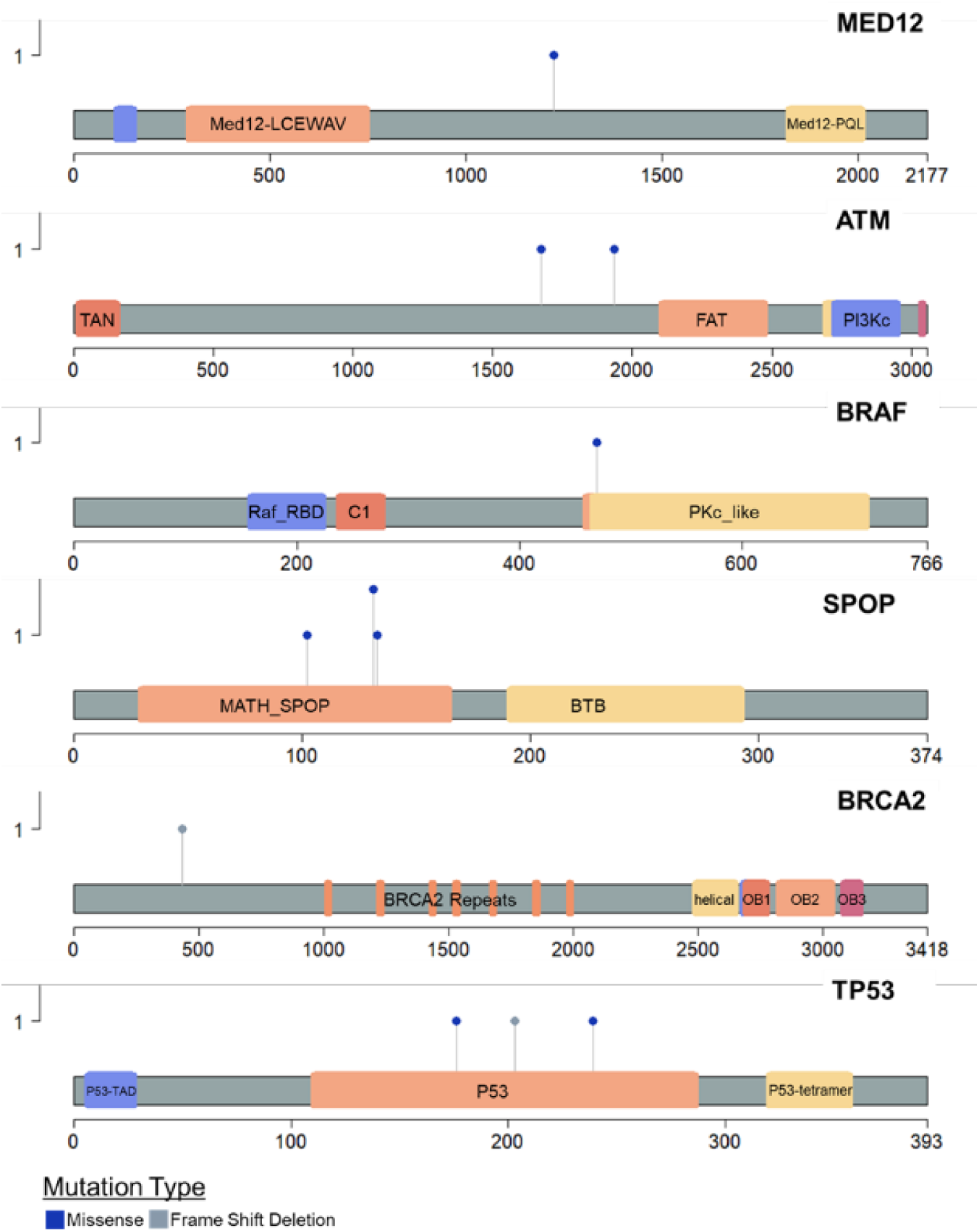
TCGA EA Somatic Variant Lollipop Plots. After filtering, the European cohort (n = 50) contained 21,957 variants. Comparison of variants with most frequently mutated NG variants showed no discernable pattern.

**Supplemental Figure 10.**
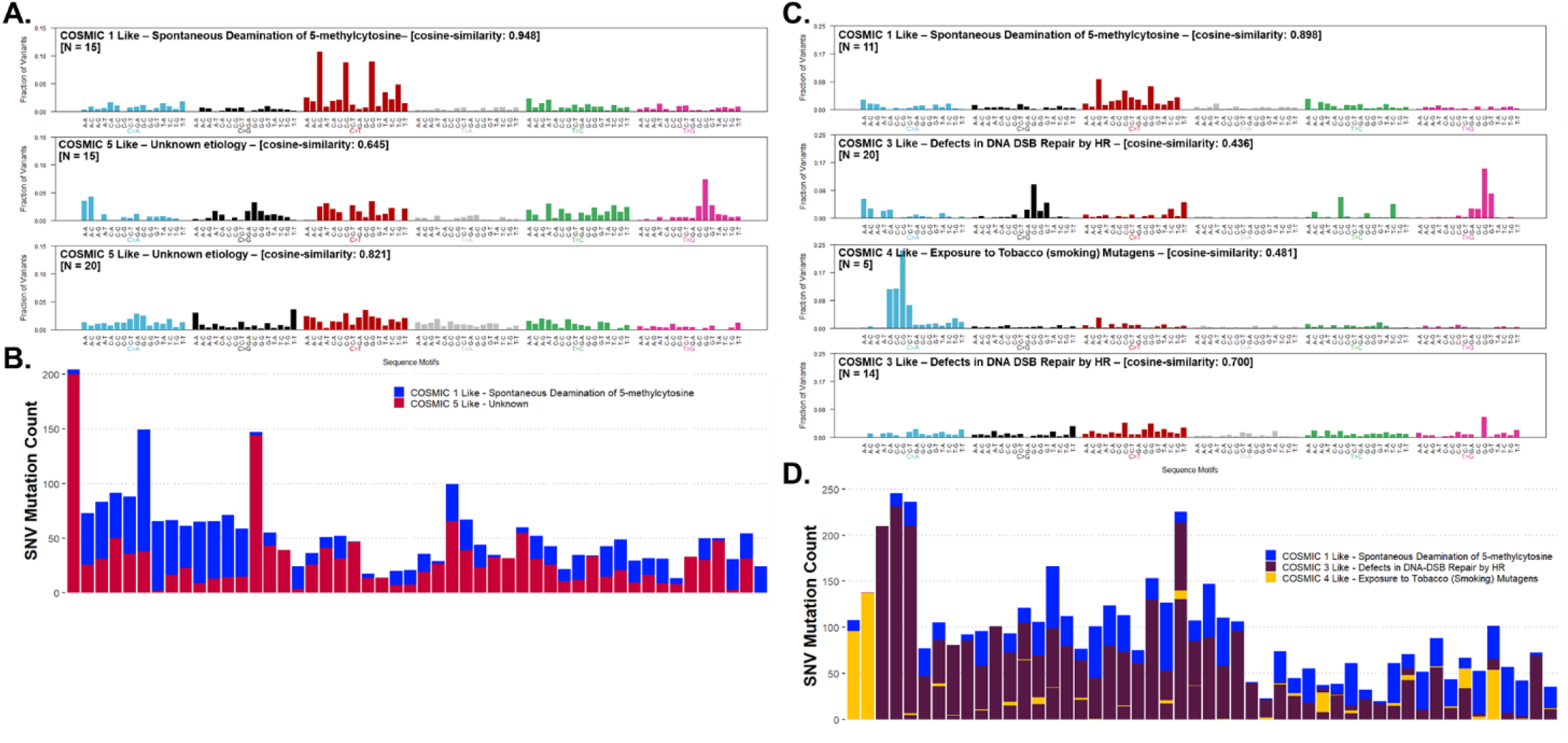
TCGA PCa Somatic Variant COSMIC Signature Analysis. Single nucleotide variations in both TCGA PCa cohorts were compared to known cancer-related mutation signatures within the Catalogue of Somatic Mutations in Cancer (COSMIC). **A and B)** TCGA AA PCa mutation patterns shared similarities (cosign similarities ≥ 0.645) with COSMIC signatures 1 and 5. **C and D)** TCGA EA PCa mutation patterns shared similarities (cosign similarities ≥ 0.436) with COSMIC signatures 1, 3, and 4.

**Supplemental Figure 11.**
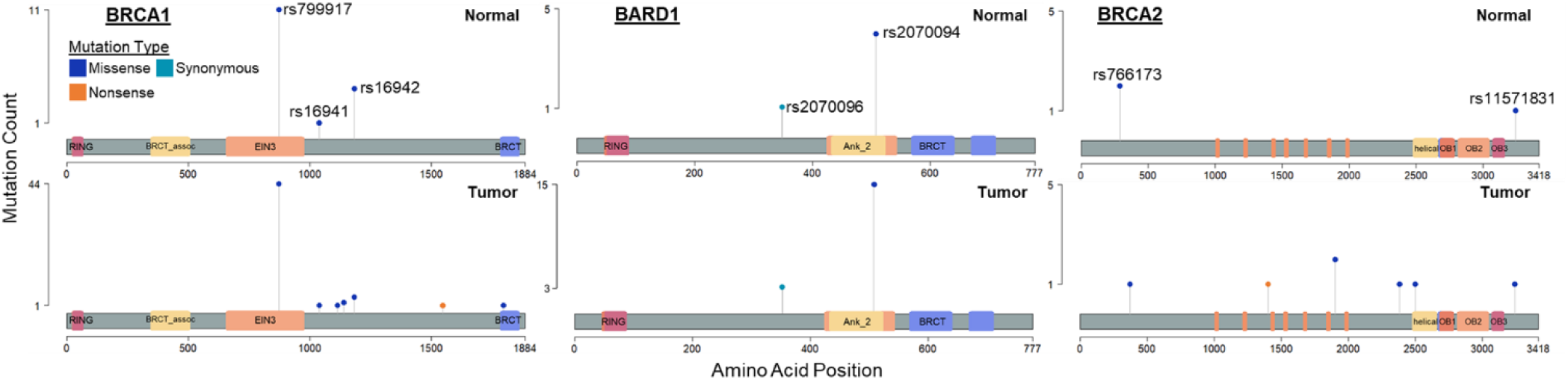
NG Germline Variant – Normal/Tumor Comparison Lollipop Plots. To validate germline variants identified within NG Normal (n=11) samples, NG tumor (n=45) samples were processed using the same pipeline and surveyed for variant frequencies in *BRCA1, BARD1*, and *BRCA2*. NG tumor samples, like NG normal samples, showed a high frequency (100%) of rs799917, moderate frequencies (30-36%) of rs2070094 and rs16942, and low frequencies (9-2%) of rs16941, rs2070096, and rs11571831. rs766173 was not present in NG tumor samples.

## References

1 Ferlay, J. et al. Estimating the global cancer incidence and mortality in 2018: GLOBOCAN sources and methods. Int J Cancer 144, 1941–1953, doi:10.1002/ijc.31937 (2019).

2 Sung, H. et al. Global Cancer Statistics 2020: GLOBOCAN Estimates of Incidence and Mortality Worldwide for 36 Cancers in 185 Countries. CA Cancer J Clin 71, 209–249, doi:10.3322/caac.21660 (2021).

3 Jensen, O. M. Cancer registration: principles and methods. Vol. 95 (IARC, 1991).

4 Pietro, G. D., Chornokur, G., Kumar, N. B., Davis, C. & Park, J. Y. Racial Differences in the Diagnosis and Treatment of Prostate Cancer. Int Neurourol J 20, S112–119, doi:10.5213/inj.1632722.361 (2016).

5 Chornokur, G., Dalton, K., Borysova, M. E. & Kumar, N. B. Disparities at presentation, diagnosis, treatment, and survival in African American men, affected by prostate cancer. Prostate 71, 985–997, doi:10.1002/pros.21314 (2011).

6 Hayes, V. M., Jaratlerdsiri, W. & Bornman, M. S. R. Prostate cancer genomics and racial health disparity. Oncotarget 9, 36650–36651, doi:10.18632/oncotarget.26399 (2018).

7 Grizzle, W. E. et al. Self-Identified African Americans and prostate cancer risk: West African genetic ancestry is associated with prostate cancer diagnosis and with higher Gleason sum on biopsy. Cancer Med 8, 6915–6922, doi:10.1002/cam4.2434 (2019).

8 Irizarry-Ramirez, M. et al. Genetic ancestry and prostate cancer susceptibility SNPs in Puerto Rican and African American men. Prostate 77, 1118–1127, doi:10.1002/pros.23368 (2017).

9 Yuan, J. et al. Integrated Analysis of Genetic Ancestry and Genomic Alterations across Cancers. Cancer Cell 34, 549–560 e549, doi:10.1016/j.ccell.2018.08.019 (2018).

10 Oak, N. et al. Ancestry-specific predisposing germline variants in cancer. Genome Med 12, 51, doi:10.1186/s13073-020-00744-3 (2020).

11 Cancer Genome Atlas Research, N. et al. The Cancer Genome Atlas Pan-Cancer analysis project. Nat Genet 45, 1113–1120, doi:10.1038/ng.2764 (2013).

12 Patin, E. et al. Dispersals and genetic adaptation of Bantu-speaking populations in Africa and North America. Science 356, 543–546, doi:10.1126/science.aal1988 (2017).

13 Zakharia, F. et al. Characterizing the admixed African ancestry of African Americans. Genome Biol 10, R141, doi:10.1186/gb-2009-10-12-r141 (2009).

14 Salas, A., Carracedo, A., Richards, M. & Macaulay, V. Charting the ancestry of African Americans. Am J Hum Genet 77, 676–680, doi:10.1086/491675 (2005).

15 Micheletti, S. J. et al. Genetic Consequences of the Transatlantic Slave Trade in the Americas. Am J Hum Genet 107, 265–277, doi:10.1016/j.ajhg.2020.06.012 (2020).

16 Stefflova, K. et al. Dissecting the within-Africa ancestry of populations of African descent in the Americas. PLoS One 6, e14495, doi:10.1371/journal.pone.0014495 (2011).

17 Gay, N. R. et al. Impact of admixture and ancestry on eQTL analysis and GWAS colocalization in GTEx. Genome Biol 21, 233, doi:10.1186/s13059-020-02113-0 (2020).

18 Kim, M. S., Patel, K. P., Teng, A. K., Berens, A. J. & Lachance, J. Genetic disease risks can be misestimated across global populations. Genome Biol 19, 179, doi:10.1186/s13059-018-1561-7 (2018).

19 Cook, M. B. et al. A genome-wide association study of prostate cancer in West African men. Hum Genet 133, 509–521, doi:10.1007/s00439-013-1387-z (2014).

20 Du, Z. et al. Genetic risk of prostate cancer in Ugandan men. Prostate 78, 370–376, doi:10.1002/pros.23481 (2018).

21 Petersen, D. C. et al. African KhoeSan ancestry linked to high-risk prostate cancer. BMC Med Genomics 12, 82, doi:10.1186/s12920-019-0537-0 (2019).

22 Jaratlerdsiri, W. et al. Whole-Genome Sequencing Reveals Elevated Tumor Mutational Burden and Initiating Driver Mutations in African Men with Treatment-Naive, High-Risk Prostate Cancer. Cancer Res 78, 6736–6746, doi:10.1158/0008-5472.CAN-18-0254 (2018).

23 Hayes, V. M. & Bornman, M. S. R. Prostate Cancer in Southern Africa: Does Africa Hold Untapped Potential to Add Value to the Current Understanding of a Common Disease? J Glob Oncol 4, 1–7, doi:10.1200/JGO.2016.008862 (2018).

24 Yadav, S. et al. Somatic mutations in the DNA repairome in prostate cancers in African Americans and Caucasians. Oncogene 39, 4299–4311, doi:10.1038/s41388-020-1280-x (2020).

25 Beebe-Dimmer, J. L., Zuhlke, K. A., Johnson, A. M., Liesman, D. & Cooney, K. A. Rare germline mutations in African American men diagnosed with early-onset prostate cancer. Prostate 78, 321–326, doi:10.1002/pros.23464 (2018).

26 de Bono, J. et al. Olaparib for Metastatic Castration-Resistant Prostate Cancer. N Engl J Med 382, 2091–2102, doi:10.1056/NEJMoa1911440 (2020).

27 Bolger, A. M., Lohse, M. & Usadel, B. Trimmomatic: a flexible trimmer for Illumina sequence data. Bioinformatics 30, 2114–2120, doi:10.1093/bioinformatics/btu170 (2014).

28 Li, H. Aligning sequence reads, clone sequences and assembly contigs with BWA-MEM. 1303.3997v1 [q-bio.GN] (2013).

29 Li, H. et al. The Sequence Alignment/Map format and SAMtools. Bioinformatics 25, 2078–2079, doi:10.1093/bioinformatics/btp352 (2009).

30 McKenna, A. et al. The Genome Analysis Toolkit: a MapReduce framework for analyzing next-generation DNA sequencing data. Genome research 20, 1297–1303, doi:10.1101/gr.107524.110 (2010).

31 Garcia-Alcalde, F. et al. Qualimap: evaluating next-generation sequencing alignment data. Bioinformatics 28, 2678–2679, doi:10.1093/bioinformatics/bts503 (2012).

32 Isaac Subirana, H. S., Joan Vila. Building Bivariate Tables: The compareGroups Package for R. Journal of Statistical Software 57, 1–16, doi:10.18637/jss.v057.i12 (2014).

33 DePristo, M. A. et al. A framework for variation discovery and genotyping using next-generation DNA sequencing data. Nat Genet 43, 491–498, doi:10.1038/ng.806 (2011).

34 Cibulskis, K. et al. Sensitive detection of somatic point mutations in impure and heterogeneous cancer samples. Nature biotechnology 31, 213–219, doi:10.1038/nbt.2514 (2013).

35 Wang, K., Li, M. & Hakonarson, H. ANNOVAR: functional annotation of genetic variants from high-throughput sequencing data. Nucleic Acids Res 38, e164, doi:10.1093/nar/gkq603 (2010).

36 Jones, S. et al. Personalized genomic analyses for cancer mutation discovery and interpretation. Sci Transl Med 7, 283ra253, doi:10.1126/scitranslmed.aaa7161 (2015).

37 Mayakonda, A., Lin, D. C., Assenov, Y., Plass, C. & Koeffler, H. P. Maftools: efficient and comprehensive analysis of somatic variants in cancer. Genome research 28, 1747–1756, doi:10.1101/gr.239244.118 (2018).

38 Liu, X., Wu, C., Li, C. & Boerwinkle, E. dbNSFP v3.0: A One-Stop Database of Functional Predictions and Annotations for Human Nonsynonymous and Splice-Site SNVs. Human mutation 37, 235–241, doi:10.1002/humu.22932 (2016).

39 Lek, M. et al. Analysis of protein-coding genetic variation in 60,706 humans. Nature 536, 285–291, doi:10.1038/nature19057 (2016).

40 Landrum, M. J. et al. ClinVar: improving access to variant interpretations and supporting evidence. Nucleic Acids Res 46, D1062–D1067, doi:10.1093/nar/gkx1153 (2018).

41 Quinlan, A. R. & Hall, I. M. BEDTools: a flexible suite of utilities for comparing genomic features. Bioinformatics 26, 841–842, doi:10.1093/bioinformatics/btq033 (2010).

42 Van der Auwera, G. A. et al. From FastQ data to high confidence variant calls: the Genome Analysis Toolkit best practices pipeline. Curr Protoc Bioinformatics 43, 11 10 11–11 10 33, doi:10.1002/0471250953.bi1110s43 (2013).

43 Alexander, D. H., Novembre, J. & Lange, K. Fast model-based estimation of ancestry in unrelated individuals. Genome research 19, 1655–1664, doi:10.1101/gr.094052.109 (2009).

44 Genomes Project, C. et al. A global reference for human genetic variation. Nature 526, 68–74, doi:10.1038/nature15393 (2015).

45 Tate, J. G. et al. COSMIC: the Catalogue Of Somatic Mutations In Cancer. Nucleic Acids Res 47, D941–D947, doi:10.1093/nar/gky1015 (2019).

46 Shannon, P. et al. Cytoscape: a software environment for integrated models of biomolecular interaction networks. Genome research 13, 2498–2504, doi:10.1101/gr.1239303 (2003).

47 Doncheva, N. T., Morris, J. H., Gorodkin, J. & Jensen, L. J. Cytoscape StringApp: Network Analysis and Visualization of Proteomics Data. J Proteome Res 18, 623–632, doi:10.1021/acs.jproteome.8b00702 (2019).

48 Bindea, G. et al. ClueGO: a Cytoscape plug-in to decipher functionally grouped gene ontology and pathway annotation networks. Bioinformatics 25, 1091–1093, doi:10.1093/bioinformatics/btp101 (2009).

49 Collins, F. S. What we do and don’t know about ‘race’, ‘ethnicity’, genetics and health at the dawn of the genome era. Nat Genet 36, S13–15, doi:10.1038/ng1436 (2004).

50 Shraga, R. et al. Evaluating genetic ancestry and self-reported ethnicity in the context of carrier screening. BMC Genet 18, 99, doi:10.1186/s12863-017-0570-y (2017).

51 Zhang, X. et al. A functional BRCA1 coding sequence genetic variant contributes to risk of esophageal squamous cell carcinoma. Carcinogenesis 34, 2309–2313, doi:10.1093/carcin/bgt213 (2013).

52 Wang, K., Xu, L., Pan, L., Xu, K. & Li, G. The functional BRCA1 rs799917 genetic polymorphism is associated with gastric cancer risk in a Chinese Han population. Tumour Biol 36, 393–397, doi:10.1007/s13277-014-2655-9 (2015).

53 Liu, D. et al. Single nucleotide polymorphisms in breast cancer susceptibility gene 1 are associated with susceptibility to lung cancer. Oncol Lett 21, 424, doi:10.3892/ol.2021.12685 (2021).

54 Shi, M. et al. A functional BRCA1 coding sequence genetic variant contributes to prognosis of triple-negative breast cancer, especially after radiotherapy. Breast Cancer Res Treat 166, 109–116, doi:10.1007/s10549-017-4395-1 (2017).

55 Liu, X. et al. Association of three common BARD1 variants with cancer susceptibility: a system review and meta-analysis. Int J Clin Exp Med 8, 311–321 (2015).

56 Koboldt, D. C. Best practices for variant calling in clinical sequencing. Genome Med 12, 91, doi:10.1186/s13073-020-00791-w (2020).

57 Koga, Y. et al. Genomic Profiling of Prostate Cancers from Men with African and European Ancestry. Clin Cancer Res 26, 4651–4660, doi:10.1158/1078-0432.CCR-19-4112 (2020).

58 Tan, H., Bao, J. & Zhou, X. Genome-wide mutational spectra analysis reveals significant cancer-specific heterogeneity. Sci Rep 5, 12566, doi:10.1038/srep12566 (2015).

59 Lawrence, M. S. et al. Mutational heterogeneity in cancer and the search for new cancer-associated genes. Nature 499, 214–218, doi:10.1038/nature12213 (2013).

60 Greenman, C. et al. Patterns of somatic mutation in human cancer genomes. Nature 446, 153–158, doi:10.1038/nature05610 (2007).

61 Cannataro, V. L., Gaffney, S. G. & Townsend, J. P. Effect Sizes of Somatic Mutations in Cancer. J Natl Cancer Inst 110, 1171–1177, doi:10.1093/jnci/djy168 (2018).

62 Brown, J. S., O’Carrigan, B., Jackson, S. P. & Yap, T. A. Targeting DNA Repair in Cancer: Beyond PARP Inhibitors. Cancer Discov 7, 20–37, doi:10.1158/2159-8290.CD-16-0860 (2017).

63 Qin, T. T. et al. Association between BRCA1 rs799917 polymorphism and breast cancer risk: A meta-analysis of 19,878 subjects. Biomed Pharmacother 68, 905–910, doi:10.1016/j.biopha.2014.08.006 (2014).

64 Yang, M., Du, X., Zhang, F. & Yuan, S. Association between BRCA1 polymorphisms rs799917 and rs1799966 and breast cancer risk: a meta-analysis. J Int Med Res 47, 1409–1416, doi:10.1177/0300060519826819 (2019).

65 Xu, G. P. et al. The association between BRCA1 gene polymorphism and cancer risk: a meta-analysis. Oncotarget 9, 8681–8694, doi:10.18632/oncotarget.24064 (2018).

66 Nicoloso, M. S. et al. Single-nucleotide polymorphisms inside microRNA target sites influence tumor susceptibility. Cancer Res 70, 2789–2798, doi:10.1158/0008-5472.CAN-09-3541 (2010).

67 Grindedal, E. M. et al. Germ-line mutations in mismatch repair genes associated with prostate cancer. Cancer Epidemiol Biomarkers Prev 18, 2460–2467, doi:10.1158/1055-9965.EPI-09-0058 (2009).

68 Castro, E. et al. Germline BRCA mutations are associated with higher risk of nodal involvement, distant metastasis, and poor survival outcomes in prostate cancer. J Clin Oncol 31, 1748–1757, doi:10.1200/JCO.2012.43.1882 (2013).

69 Ledet, E. M. et al. Comparison of germline mutations in African American and Caucasian men with metastatic prostate cancer. Prostate 81, 433–439, doi:10.1002/pros.24123 (2021).

70 Pitt, J. J. et al. Characterization of Nigerian breast cancer reveals prevalent homologous recombination deficiency and aggressive molecular features. Nat Commun 9, 4181, doi:10.1038/s41467-018-06616-0 (2018).

71 Fackenthal, J. D. et al. High prevalence of BRCA1 and BRCA2 mutations in unselected Nigerian breast cancer patients. Int J Cancer 131, 1114–1123, doi:10.1002/ijc.27326 (2012).

72 Ricks-Santi, L. et al. Next Generation Sequencing Reveals High Prevalence of BRCA1 and BRCA2 Variants of Unknown Significance in Early-Onset Breast Cancer in African American Women. Ethn Dis 27, 169–178, doi:10.18865/ed.27.2.169 (2017).

73 Marshall, C. H. et al. Prevalence of DNA repair gene mutations in localized prostate cancer according to clinical and pathologic features: association of Gleason score and tumor stage. Prostate Cancer Prostatic Dis 22, 59–65, doi:10.1038/s41391-018-0086-1 (2019).

74 Prakash, R., Zhang, Y., Feng, W. & Jasin, M. Homologous recombination and human health: the roles of BRCA1, BRCA2, and associated proteins. Cold Spring Harb Perspect Biol 7, a016600, doi:10.1101/cshperspect.a016600 (2015).

75 Nombela, P. et al. BRCA2 and Other DDR Genes in Prostate Cancer. Cancers (Basel) 11, doi:10.3390/cancers11030352 (2019).

76 Jeggo, P. A. & Lobrich, M. How cancer cells hijack DNA double-strand break repair pathways to gain genomic instability. Biochem J 471, 1–11, doi:10.1042/BJ20150582 (2015).

77 Choi, M., Kipps, T. & Kurzrock, R. ATM Mutations in Cancer: Therapeutic Implications. Mol Cancer Ther 15, 1781–1791, doi:10.1158/1535-7163.MCT-15-0945 (2016).

78 Ma, J. et al. Genomic analysis of exceptional responders to radiotherapy reveals somatic mutations in ATM. Oncotarget 8, 10312–10323, doi:10.18632/oncotarget.14400 (2017).

79 Gao, B. et al. Functional properties of a new voltage-dependent calcium channel alpha(2)delta auxiliary subunit gene (CACNA2D2). J Biol Chem 275, 12237–12242, doi:10.1074/jbc.275.16.12237 (2000).

80 Fiske, J. L., Fomin, V. P., Brown, M. L., Duncan, R. L. & Sikes, R. A. Voltage-sensitive ion channels and cancer. Cancer Metastasis Rev 25, 493–500, doi:10.1007/s10555-006-9017-z (2006).

81 Warnier, M. et al. CACNA2D2 promotes tumorigenesis by stimulating cell proliferation and angiogenesis. Oncogene 34, 5383–5394, doi:10.1038/onc.2014.467 (2015).

82 Mellad, J. A., Warren, D. T. & Shanahan, C. M. Nesprins LINC the nucleus and cytoskeleton. Curr Opin Cell Biol 23, 47–54, doi:10.1016/j.ceb.2010.11.006 (2011).

83 Chow, K. H., Factor, R. E. & Ullman, K. S. The nuclear envelope environment and its cancer connections. Nat Rev Cancer 12, 196–209, doi:10.1038/nrc3219 (2012).

84 Sur-Erdem, I. et al. Nesprin-1 impact on tumorigenic cell phenotypes. Mol Biol Rep 47, 921–934, doi:10.1007/s11033-019-05184-w (2020).

85 Tang, B.L. ADAMTS: a novel family of extracellular matrix proteases. Int J Biochem Cell Biol 33, 33–44, doi:10.1016/s1357-2725(00)00061-3 (2001).

86 Discher, D. E. et al. Matrix Mechanosensing: From Scaling Concepts in ‘Omics Data to Mechanisms in the Nucleus, Regeneration, and Cancer. Annu Rev Biophys 46, 295–315, doi:10.1146/annurev-biophys-062215-011206 (2017).

87 Egeblad, M., Rasch, M. G. & Weaver, V. M. Dynamic interplay between the collagen scaffold and tumor evolution. Curr Opin Cell Biol 22, 697–706, doi:10.1016/j.ceb.2010.08.015 (2010).

88 Caswell-Jin, J. L. et al. Racial/ethnic differences in multiple-gene sequencing results for hereditary cancer risk. Genet Med 20, 234–239, doi:10.1038/gim.2017.96 (2018).

89 Ndugga-Kabuye, M. K. & Issaka, R. B. Inequities in multi-gene hereditary cancer testing: lower diagnostic yield and higher VUS rate in individuals who identify as Hispanic, African or Asian and Pacific Islander as compared to European. Fam Cancer 18, 465–469, doi:10.1007/s10689-019-00144-6 (2019).

90 Saulsberry, K. & Terry, S. F. The need to build trust: a perspective on disparities in genetic testing. Genet Test Mol Biomarkers 17, 647–648, doi:10.1089/gtmb.2013.1548 (2013).

91. Tan, S. H., Petrovics, G. & Srivastava, S. Prostate cancer genomics: Recent advances and the prevailing underrepresentation from racial and ethnic minorities. International Journal of Molecular Sciences (2018) doi:10.3390/ijms19041255.

